# Large Language Models for Psychiatric Phenotype Extraction from Electronic Health Records

**DOI:** 10.1101/2025.08.07.25333172

**Authors:** Clara Frydman-Gani, Alejandro Arias, Maria Perez Vallejo, John Daniel Londoño Martínez, Johanna Valencia-Echeverry, Mauricio Castaño, Alex A. T. Bui, Nelson B. Freimer, Carlos Lopez-Jaramillo, Loes M. Olde Loohuis

## Abstract

The accurate detection of clinical phenotypes from electronic health records (EHRs) is pivotal for advancing large-scale genetic and longitudinal studies in psychiatry. Free-text clinical notes are an essential source of symptom-level information, particularly in psychiatry. However, the automated extraction of symptoms from clinical text remains challenging.

Here, we tested 11 open-source generative large language models (LLMs) for their ability to detect 109 psychiatric phenotypes from clinical text, using annotated EHR notes from a psychiatric clinic in Colombia. The LLMs were evaluated both “out-of-the-box” and after fine-tuning, and compared against a traditional natural language processing (tNLP) method developed from the same data. We show that while base LLM performance was poor to moderate (0.2-0.6 macro-F1 for zero-shot; 0.2-0.74 macro-F1 for few shot), it improved significantly after fine-tuning (0.75-0.86 macro-F1), with several fine-tuned LLMs outperforming the tNLP method. In total, 100 phenotypes could be reliably detected (F1>0.8) using either a fine-tuned LLM or tNLP.

To generate a fine-tuned LLM that can be shared with the scientific and medical community, we created a fully synthetic dataset free of patient information but based on original annotations. We fine-tuned a top-performing LLM on this data, creating “Mistral-small-psych”, an LLM that can detect psychiatric phenotypes from Spanish text with performance comparable to that of LLMs trained on real EHR data (macro-F1=0.79).

Finally, the fine-tuned LLMs underwent an external validation using data from a large psychiatric hospital in Colombia, the Hospital Mental de Antioquia, highlighting that most LLMs generalized well (0.02-0.16 point loss in macro-F1). Our study underscores the value of domain-specific adaptation of LLMs and introduces a new model for accurate psychiatric phenotyping in Spanish text, paving the way for global precision psychiatry.

## Introduction

The large-scale extraction of psychiatric phenotypes from electronic health records (EHRs) enables epidemiological, translational, and genetic studies that transcend the boundaries of traditional diagnostic categories of severe mental illnesses (SMIs)^1–3^. Moreover, fine-grained psychiatric symptoms and social determinants of health can be used as features in machine learning models for predicting clinical outcomes, such as diagnostic conversion, risk of relapse, suicidality, and treatment response^4–7^. Free text notes within the EHR are a valuable source of such symptom-level information, particularly in psychiatry, where the absence of biomarkers means most phenotypes are recorded through patient self-reports and clinicians’ narrative observations.

To this end, natural language processing (NLP) techniques, and specifically large language models (LLMs), are an exciting, quickly-evolving frontier enabling the automated and scalable extraction of phenotypes from clinical text. Generative LLMs are neural network-based NLP systems based on the Transformer architecture^8^. Applications of such models rely on transfer-learning to complete a wide range of downstream tasks, and their performance for domain-specific uses can be further refined using in-context learning or fine-tuning. Generative LLMs have demonstrated impressive performance in various clinical applications^9–12^.

LLMs have been applied to automate the extraction of clinical concepts, phenotypes, and other health-related entities from English-language clinical text^13–15^. While some studies have begun to explore the potential of LLMs in extracting psychiatric phenotypes from text, current efforts have several limitations. First, existing efforts typically target a small set of features^16–19^, or focus on one or few, typically encoder-based LLMs^20–22^. Second, there is limited availability of large-scale, high-quality annotated EHR data^3,23^, which is essential for fine-tuning and accurate evaluation of model performance. Given this limitation, many studies rely on non-EHR text sources, such as social media and instant messaging platforms^24–26^, potentially limiting their applicability to clinical settings; others use clinical data from a single source^27^, and are unable to test generalizability across settings. Third, the application of closed-source generative LLMs on EHR-derived text may raise concerns around patient privacy and potential leakage of protected health information^23^. This constraint has motivated the use of open-source LLMs^28,29^, which, while applicable to EHR data, cannot be publicly released after fine-tuning due to their risk of leaking identifiable data. Fourth, while recent work has begun to explore the utility of NLP for targeting certain clinical^30,31^ and psychiatric^32^ concepts in Spanish-language text, the phenotyping capabilities of generative LLMs in Spanish texts remain understudied. This represents a critical gap, given that Spanish is the world’s second most common native language, and is widely used in clinical practice across multiple countries. Nevertheless, the development of clinical NLP tools that could serve large Spanish-speaking populations remains limited.

To address these gaps, we evaluated the performance of 11 state-of-the-art open-source generative LLMs for the extraction of 136 psychiatric phenotypes from Spanish-language EHR documents, including symptoms related to mood and affect, substance use, perception, and behavior. Models were applied using both zero and few-shot prompting, with the latter providing a small number of examples taken from annotated documents during inference. Next, we fine-tuned all LLMs using a clinician-annotated corpus of EHR documents, and compared their performance to that of a pattern-based traditional NLP (tNLP) method previously developed in the same setting^33^. We then developed Mistral-small-psych, a new open-source light-weight generative LLM that circumvents the data leakage limitation of LLMs by being trained on synthetic data generated using real phenotype annotations. Finally, we evaluated all fine-tuned models on two EHR-derived test sets to assess performance and generalizability across hospital settings.

## Methods

### Models, Settings and Data

#### Model Selection

We selected an array of widely-used generative LLMs of varying sizes for benchmarking and fine-tuning. Two key criteria in model selection were pretraining on corpora that included Spanish-language text, and open-source availability. The latter requirement was necessary given that the evaluation and fine-tuning data were derived from EHRs and had to be contained in a secure environment, requiring the ability to deploy and run all models locally.

The following models were selected for this study: Llama3-8B-Instruct (hereafter: “Llama3-8B”) and Llama3-70B-Instruct (“Llama3-70B”) released by Meta AI^34^, Qwen2.5-7B-Instruct (“Qwen2.5-7B”), Qwen2.5-14B-Instruct (“Qwen2.5-14B”), and Qwen2.5-32B-Instruct (“Qwen2.5-32B”) by Alibaba Cloud^35^, Mistral-7B-Instruct-v0.3 (“Mistral-7B”), Mistral-small-24B-Instruct-2501 (“Mistral-small”) and Mixtral-8x7B-Instruct-v0.1 (“Mixtral-8x7B”) by Mistral AI^36,37^, Mistral-nemo-Instruct-2407 (“Mistral-nemo”) by Mistral AI and NVIDIA^38^, and aya-23-8B (“aya-8B”) and aya-23-35B (“aya-35B”) by Cohere^39^. An overview of the selected LLMs is shown in Table 1.

**Table 1:**
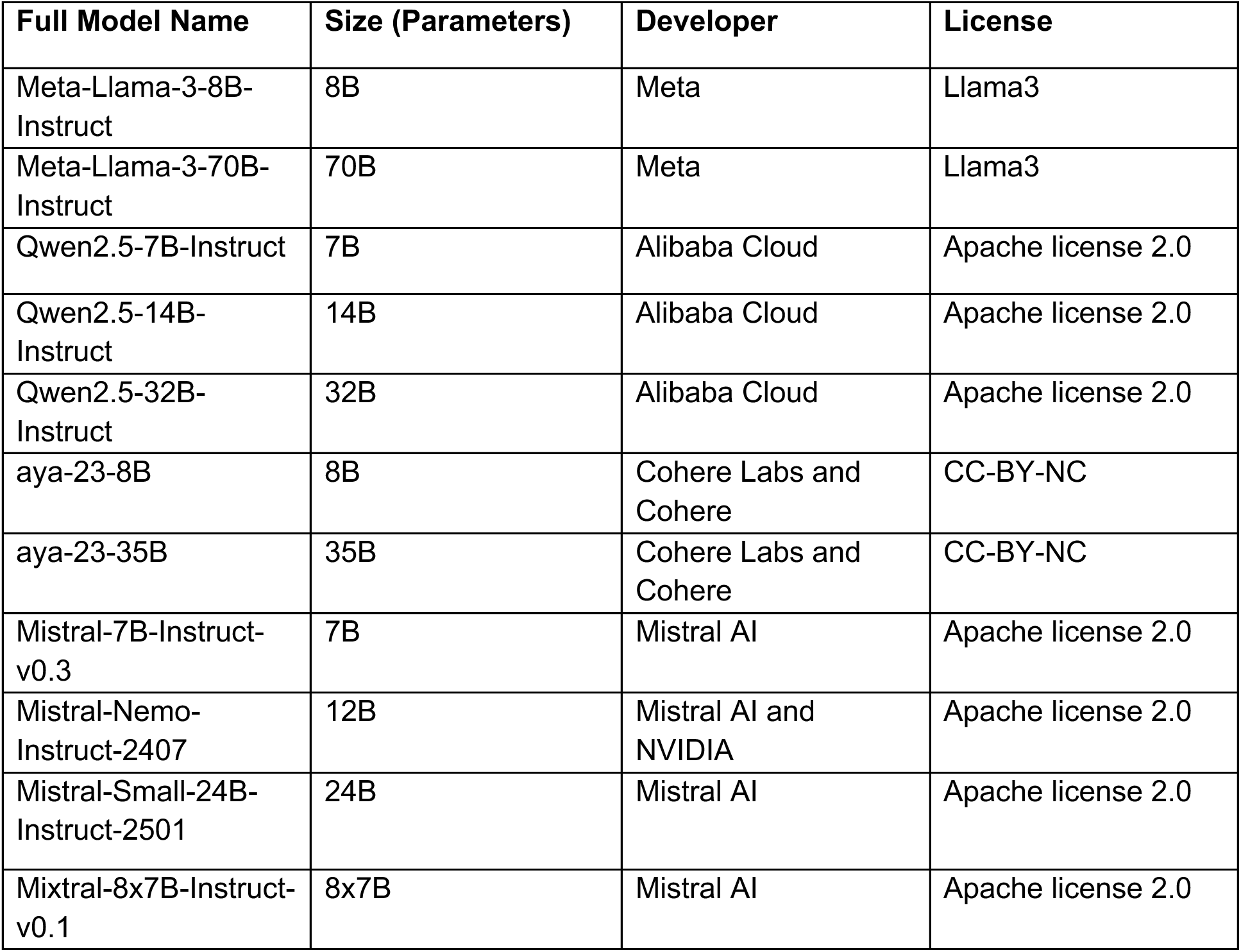
Model information.

#### Setting

We used data from two distinct settings in the model evaluation and fine-tuning processes: the first, Clínica San Juan de Dios Manizales (CSJDM) in Manizales, Colombia, serves a population of ∼1 million individuals in the department of Caldas. The second, Hospital Mental de Antioquia (HOMO), is a larger psychiatric institution and the only public psychiatric hospital in the department of Antioquia, Colombia. Data from CSJDM was used to evaluate and fine-tune all LLMs, as well as for the construction of a synthetic dataset, while data from HOMO was subsequently used as an out-of-domain test set to evaluate the generalizability of all fine-tuned models.

#### Real EHR Data and Phenotype Annotation

We used a previously curated corpus of 2,000 clinical documents from the CSJDM clinical notes^33^. Documents were selected, annotated, and split into training and test sets as described in^33^. Briefly, documents were selected from intake, emergency department, and outpatient notes, prioritizing concept-rich fields. Then, two expert clinicians – a psychiatrist and a clinical psychologist - annotated the documents for over 130 phenotypes, including phenotypes relating to mood/affect (such as “guilt”, “hopelessness”, and “depressed mood”), thought and cognition (“delusions”, “circumstantiality”, and “suicidal ideation”), and behavior and motor function (“aggression”, “disorganized behavior”), among others (Figure 1A, see Supplementary Table 1 for a list of all phenotypes). Documents were split into training and test sets (82%-18%). Test set documents (n=358 documents) underwent a joint revision process resolving any discrepancies among the annotators to ensure a rigorous reliability standard for evaluation. For the fine-tuning step, the initial CSJDM training set (n=1,642) was split into training (n=1,477) and validation (n=165) sets. The same process was applied to HOMO EHR documents to generate the HOMO test set, consisting of 309 documents.

**Figure 1:**
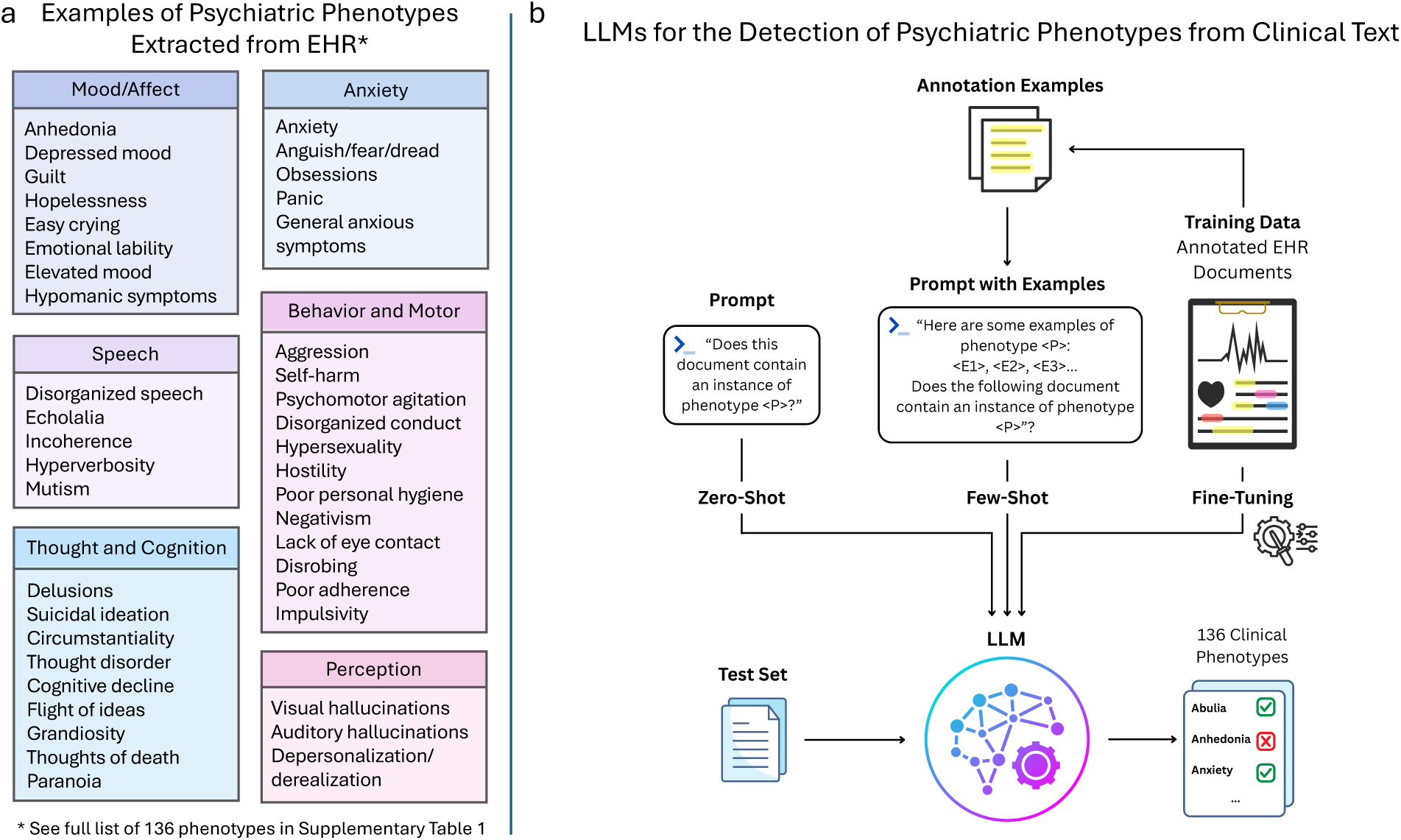
Overview of extracted phenotypes and study design. (a) Examples of psychiatric phenotypes extracted from EHR in this study (the full list can be found in Supplementary Table 1). (b) LLM application overview: LLMs were evaluated on the detection of 136 psychiatric phenotypes from EHR documents in zero- and few-shot settings (with the latter including up to five annotated examples of the phenotype of interest in the prompt); additionally, LLMs were fine-tuned using a training set of 1,477 annotated EHR documents.

#### Synthetic Data

To enable model fine-tuning and publication without risk of protected health data leakage, we created a synthetic dataset of documents not based on real clinical text. To do so, we prompted Llama3-70B to generate sentences emulating clinical notes written by a clinician, containing real CSJDM EHR training set annotations (Supplementary Note 1). To ensure data privacy, the LLM was prompted using only the specific annotated spans – previously published brief phrases, manually reviewed to ensure no identifiable information was included. As such, the model was at no point exposed to patient information, and the generated sentences carried no risk of containing it. Additionally, seeing that the real CSJDM data included many sentences with mostly somatic information and no mention of psychiatric symptoms (such as laboratory test results and depiction of somatic symptoms), we also prompted Llama3-70B to generate clinical sentences with no mentions of psychiatric symptoms (hence referred to as “negative sentences”, Supplementary Note 1).

Synthetic sentences underwent a quality control step ensuring the inclusion of the intended annotations (Supplementary Note 1). Then, for each phenotype, we sampled 3,440 sentences while prioritizing diverse annotation examples, as well as 300 random negative sentences, for a total of 3,740 synthetic sentences (Supplementary Note 1). Finally, sentences were shuffled and assembled into groups of 1 to 6 sentences to produce longer “documents”.

As the LLM was prompted to generate synthetic sentences intended to emulate clinicians’ clinical notes, the synthetic data often contained phrases depicting additional mentions of various psychiatric phenotypes, other than the one the LLM was explicitly prompted for. To recover and label these additional symptoms incidentally appearing in the synthetic dataset, and create more accurately labeled training data, we applied the tNLP method to the synthetic documents as a weak-labeling approach. The final synthetic training set contained annotations for both the originally targeted phenotypes and those identified by the tNLP method.

### Inference Applications

#### Zero-Shot Prompting

All LLMs were evaluated on the CSJDM test set, prompted to provide a binary response indicating the presence of a given phenotype in each test set document (Figure 1B, see Supplementary Note 2 for full detail). For this evaluation, we used zero-shot prompting (i.e., prompting the models to extract psychiatric symptoms providing no examples or additional information). LLMs were prompted separately for each document-phenotype pair, with no shared context between prompts, ensuring that no information was carried over across queries. All LLMs were served via Ollama v0.3.14, and loaded in 4-bit quantization on instances utilizing one or four NVIDIA A10G Tensor Core GPUs, with 24 or 96 GB of VRAM, respectively.

#### Few-Shot Prompting

Few-shot prompting consists of supplying a small number of examples of the task (typically ranging from 2-10) as part of the prompt. This approach leverages in-context learning, wherein the model conditions its response on the examples and information provided when prompting, with no modification to its underlying weights; it has been demonstrated to improve model performance, in some cases yielding comparable results to those of model fine-tuning, but at a fraction of the necessary labeled data and computational cost^40^.

Here, we selected up to five examples of annotated spans from the CSJDM training set for each phenotype while ensuring a balance of example diversity and representativeness and included them in the prompt as examples (Figure 1B, Supplementary Note 3). As before, each combination of test set document-phenotype was prompted separately, with no context carried between prompts.

### Fine-Tuning

#### Fine-Tuning on CSJDM Data

We utilized the CSJDM training set to fine-tune all LLMs (Figure 1B). LLMs were trained for 1-4 epochs on G5 AWS instances with 1 (Llama3-8B, Mistral-7B, Mistral-nemo, Mistral-small, Qwen2.5-7B, Qwen2.5-14B, Qwen2.5-32B, aya-8B) or 4 (aya-35B, Mixtral-8x7B, Llama3-70B) NVIDIA A10G Tensor Core GPUs, with 24GB of VRAM each. To optimize memory use and training time, we used Unsloth^41^ in the fine-tuning of Llama3-8B, Mistral-7B, Mistral-nemo, Mistral-small, Qwen2.5-7B, Qwen2.5-14B, and Qwen2.5-32B models, and Axolotl for all other models. To further reduce memory and training time requirements, all models were downloaded in 4-bit quantization, and fine-tuned using Low-Rank Adaptation (LoRA), a parameter-efficient fine-tuning (PEFT) method in which the original model’s weights remain frozen, while smaller rank decomposition matrices injected into the model’s layers are trained during fine-tuning^42^.

LoRA is a widely used method in LLM fine-tuning, enabling a significant reduction in the number of trainable parameters without significant losses in performance. To address the limitations of LLM output explainability, we utilized an instruction-tuning approach in which the models were trained to produce not only the phenotypes detected in the text, but also their corresponding labeled spans. Additional information on the fine-tuning process can be found in Supplementary Note 4.

#### Fine-Tuning on Synthetic Data

The top-performing base model from the real-data fine-tuning step - prioritizing smaller model size in cases of similar performance, to enable greater accessibility - was fine-tuned using the synthetic dataset. As before, fine-tuning was performed using instruction-tuning requiring outputs to contain the detected phenotypes as well as their labeled spans, with a 4-bit quantized base model, using Unsloth and LoRA (Supplementary Note 5).

### Evaluation

#### Model Performance Metrics

Performance at all levels (for both inference and fine-tuned models) was evaluated on the CSJDM test set against the clinicians’ annotations, using precision, recall, macro-F1, and accuracy (Supplementary Note 6).

To ensure a minimum level of reliability in performance metrics, we report performance only for phenotypes with at least three instances in the CSJDM training and test sets (N phenotypes=109).

For interpretation, overall model performance based on macro-F1 scores was considered poor (<=0.6), moderate (0.6-0.75), good (0.75-0.9) or excellent (>=0.9).

#### Comparison to tNLP Method

LLM performance (both for inference and after fine-tuning) was benchmarked against a tNLP method previously developed from the same CSJDM training data^33^ and evaluated on the CSJDM test set. Performance comparisons of this method with the LLMs are based on the 88 phenotypes evaluated using tNLP. Overall model performances were compared using a Wilcoxon signed-rank test comparing F1 scores, using FDR correction for multiple testing with an alpha value of 0.05.

#### Cross-Hospital Transportability Evaluation

All fine-tuned models underwent subsequent evaluation on the HOMO out-of-domain test set, to assess their performance in external settings.

### Dataset Size Sensitivity Analysis

To assess how model performance changes as a function of the synthetic training dataset size, we created multiple training subsets of increasing sizes from the original synthetic training data by randomly sampling three subsets of 250, 500, 750 and 1000 synthetic documents. Separate models were fine-tuned on each dataset, and their performance was evaluated on the CSJDM test set.

### Inference Runtime, Electricity Consumption and Cost

The application of generative LLMs for inference typically requires the utilization of specialized hardware (i.e., graphical processing units, or GPUs). These applications could incur substantial costs, time, and carbon footprint when applied to real-world EHR repositories, which can consist of millions of documents. To benchmark these expenses and compare the relative resource requirements of the models evaluated here, we report the average runtime (in seconds), system electricity consumption (in watt-hours) and cost (in cents) for running zero-shot inference with each LLM via Ollama on a single phenotype across 100 documents. Consumption measurements were performed using Code Carbon^43^. While some LLMs could run on smaller hardware, all measurements were standardized on an AWS G5.12xlarge EC2 instance for consistency. Costs were estimated based on runtime and AWS instance pricing in the U.S. West (Oregon) Region.

## Results

### Model Performance

#### Zero-Shot Prompting

All LLMs were assessed for their capabilities to detect 109 psychiatric phenotypes associated with mood and affect, perception, thought and cognition, and behavior, among other domains (Figure 1, Supplementary Table 1). Overall, model performance was poor to moderate in zero-shot evaluations across the 109 phenotypes tested (Figure 2, Table 2). While larger models within the same family often performed better than smaller ones, this trend did not extend across different model families (Supplementary Figure 1A).

**Figure 2:**
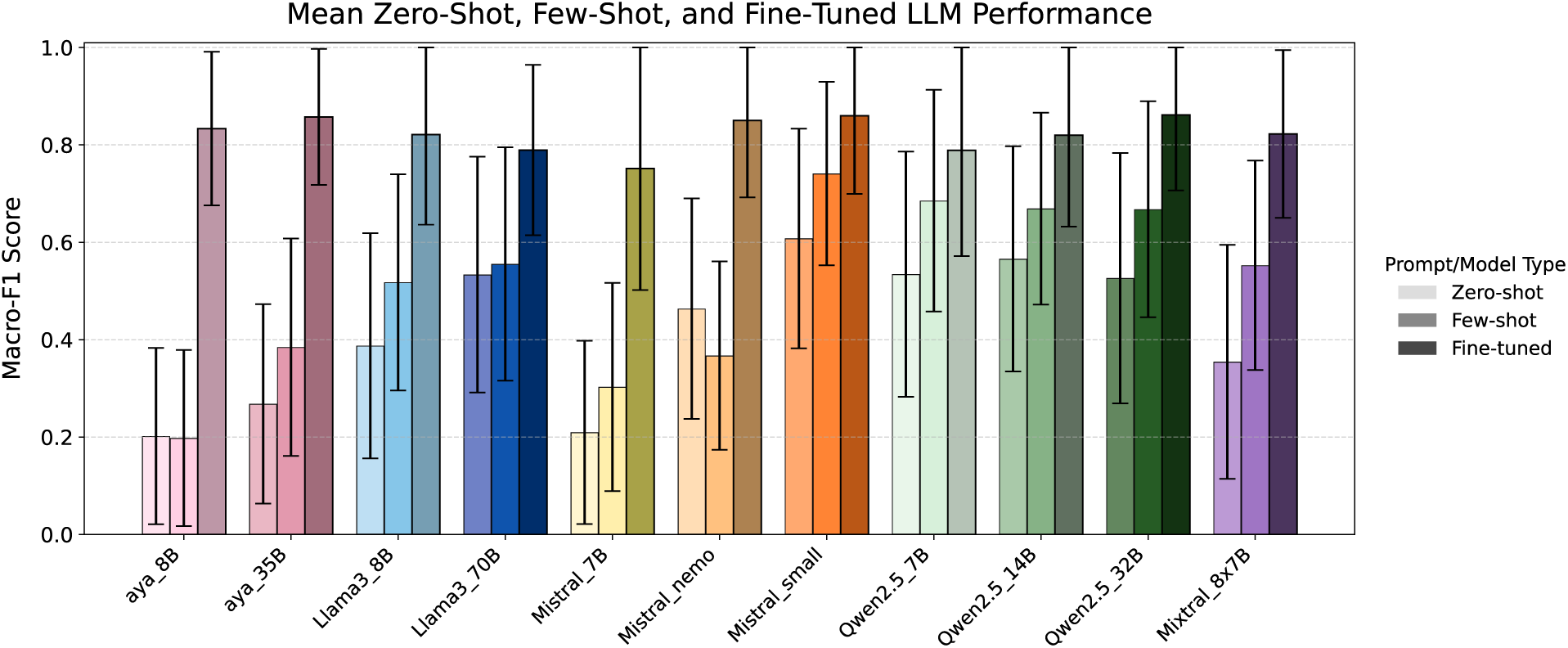
Mean performance (macro-F1) for all models. Mean LLM performance (macro-F1) on the CSJDM test set (n=358 documents), in zero-shot (light bars, left) and few-shot (medium bars, middle) settings, as well as after fine-tuning (dark bars, right) for all testable phenotypes (n=109 phenotypes). Error bars represent a standard deviation from the mean F1 score.

**Table 2:**
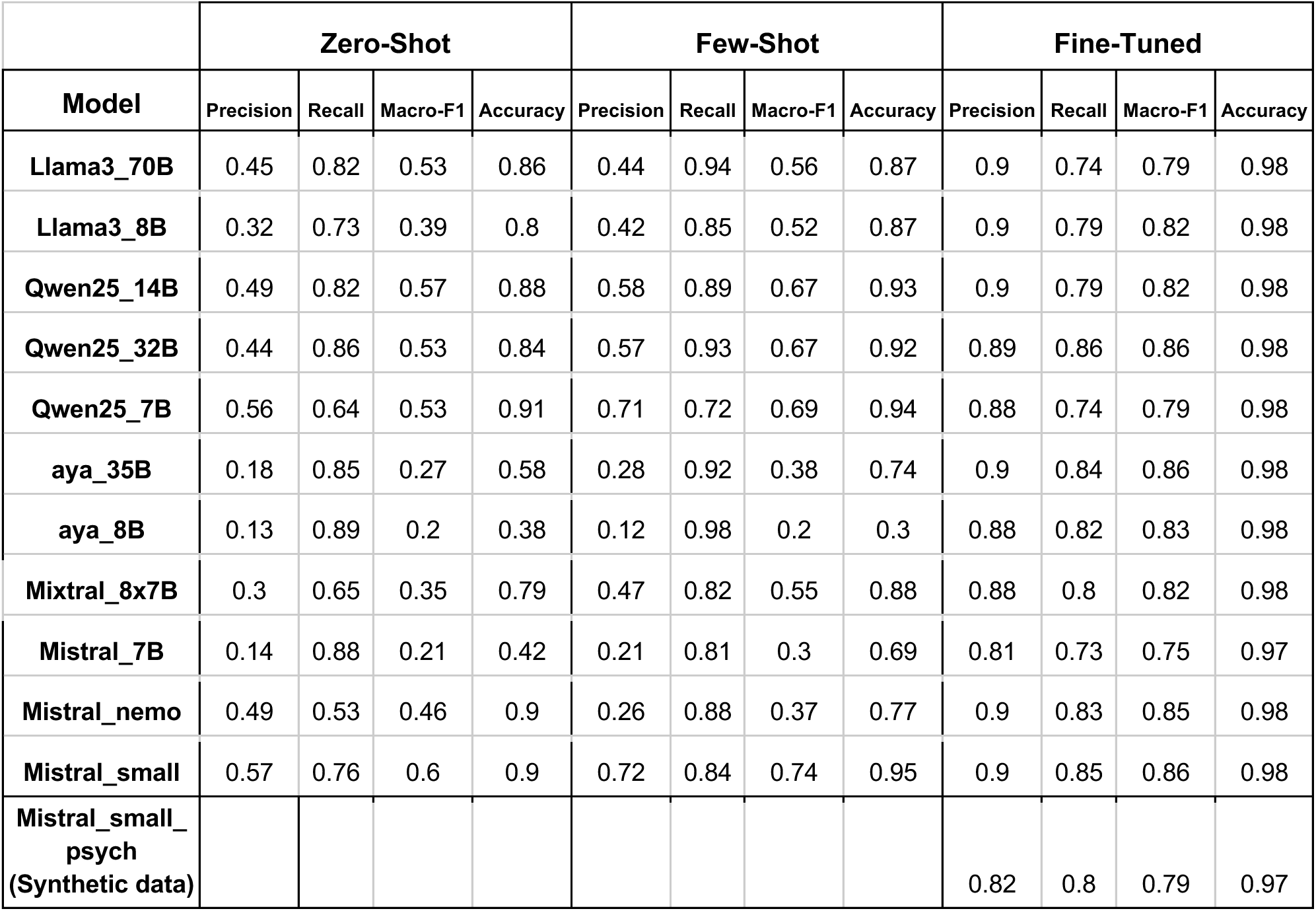
Zero-shot, few-shot and fine-tuned performance metrics (precision, recall, macro-F1, accuracy) for all LLMs.

|  | Zero-Shot |  |  |  | Few-Shot |  |  |  | Fine-Tuned |  |  |  |
| --- | --- | --- | --- | --- | --- | --- | --- | --- | --- | --- | --- | --- |
| Model | Precision | Recall | Macro-F1 | Accuracy | Precision | Recall | Macro-F1 | Accuracy | Precision | Recall | Macro-F1 | Accuracy |
| Llama3_70B | 0.45 | 0.82 | 0.53 | 0.86 | 0.44 | 0.94 | 0.56 | 0.87 | 0.9 | 0.74 | 0.79 | 0.98 |
| Llama3_8B | 0.32 | 0.73 | 0.39 | 0.8 | 0.42 | 0.85 | 0.52 | 0.87 | 0.9 | 0.79 | 0.82 | 0.98 |
| Qwen25_14B | 0.49 | 0.82 | 0.57 | 0.88 | 0.58 | 0.89 | 0.67 | 0.93 | 0.9 | 0.79 | 0.82 | 0.98 |
| Qwen25_32B | 0.44 | 0.86 | 0.53 | 0.84 | 0.57 | 0.93 | 0.67 | 0.92 | 0.89 | 0.86 | 0.86 | 0.98 |
| Qwen25_7B | 0.56 | 0.64 | 0.53 | 0.91 | 0.71 | 0.72 | 0.69 | 0.94 | 0.88 | 0.74 | 0.79 | 0.98 |
| aya_35B | 0.18 | 0.85 | 0.27 | 0.58 | 0.28 | 0.92 | 0.38 | 0.74 | 0.9 | 0.84 | 0.86 | 0.98 |
| aya_8B | 0.13 | 0.89 | 0.2 | 0.38 | 0.12 | 0.98 | 0.2 | 0.3 | 0.88 | 0.82 | 0.83 | 0.98 |
| Mixtral_8x7B | 0.3 | 0.65 | 0.35 | 0.79 | 0.47 | 0.82 | 0.55 | 0.88 | 0.88 | 0.8 | 0.82 | 0.98 |
| Mistral_7B | 0.14 | 0.88 | 0.21 | 0.42 | 0.21 | 0.81 | 0.3 | 0.69 | 0.81 | 0.73 | 0.75 | 0.97 |
| Mistral_nemo | 0.49 | 0.53 | 0.46 | 0.9 | 0.26 | 0.88 | 0.37 | 0.77 | 0.9 | 0.83 | 0.85 | 0.98 |
| Mistral_small | 0.57 | 0.76 | 0.6 | 0.9 | 0.72 | 0.84 | 0.74 | 0.95 | 0.9 | 0.85 | 0.86 | 0.98 |
| Mistral_small_psych<br>(Synthetic data) |  |  |  |  |  |  |  |  | 0.82 | 0.8 | 0.79 | 0.97 |

Mistral-small (macro-F1=0.6, mean accuracy=0.9) and Qwen2.5-14B (macro-F1=0.57, mean accuracy=0.88) achieved the best overall zero-shot performance. However, phenotype-level performance varied substantially, reflecting the variability in the models’ capability of detecting specific phenotypes (Supplementary Figure 2A, Supplementary Table 2). While most models had strong (>=0.8) recall, they were limited by their precision. In particular, the smaller Mistral-7B and aya-8B showed good recall (0.88 and 0.89, respectively) but markedly low precision (0.14 and 0.13, respectively), suggesting a tendency toward positive labeling with limited discriminative ability in this setting. The tNLP method significantly outperformed all LLMs (Supplementary Figure 3, Supplementary Table 5), achieving a macro-F1 score of 0.85, with particularly notable advantages in some phenotypes, including “negativism” and “thought disorder” (Supplementary Figure 2B, Supplementary Table 6). In contrast, a small number of phenotypes, including “weight loss” and “poor personal hygiene”, were better detected by several LLMs.

#### Few-Shot Prompting

Few-shot prompting improved macro-F1 for all models except Mistral-nemo (-0.09) and aya-8B (no change) (Figure 2, Table 2), with phenotype-level performance shown in Supplementary Figure 4). Mistral-small achieved the highest few-shot performance (macro-F1=0.74, mean accuracy=0.95) following an increase in both precision and recall following the addition of examples to the prompt. The phenotypes with the greatest average improvement from zero-to few-shot across all LLMs were “low energy” (+0.4 F1), “hyperverbosity” (+0.37 F1), “psychomotor delay” and “poor insight” (+0.39 F1 for both, Supplementary Table 3). Despite this marked improvement for most models, the tNLP method significantly outperformed all LLMs in the few-shot setting (Supplementary Figure 3, Supplementary Tables 5,6).

#### Real-Data Fine-Tuning

All LLMs saw dramatic improvement in performance following the fine-tuning process, displaying good performance (Figure 2, Table 2, Supplementary Table 4). The best performing fine-tuned models were aya-35B, Mistral-small, and Qwen2.5-32B (macro-F1=0.86 for all). Aya-8B displayed the greatest improvement compared to the base model’s zero-shot performance, with a macro-F1 increase of +0.63.

While performance was more variable - across both models and phenotypes - in ultra-rare phenotypes, it generally improved and converged as the phenotypes’ training set frequency increased (Figure 3, Supplementary Figure 5A). Nevertheless, we still observed variation in some cases, both between models (e.g., Qwen2.5-7B’s markedly low performance for “poor adherence”) and for specific phenotypes (e.g., all models’ relatively poor performance for “negativism”, “adverse effects” and “poor response to psychotropic medications”). In contrast, some phenotypes, like “hypothymia”, had high (F1>=0.89) performance for all models despite having relatively low frequencies in the training data.

**Figure 3:**
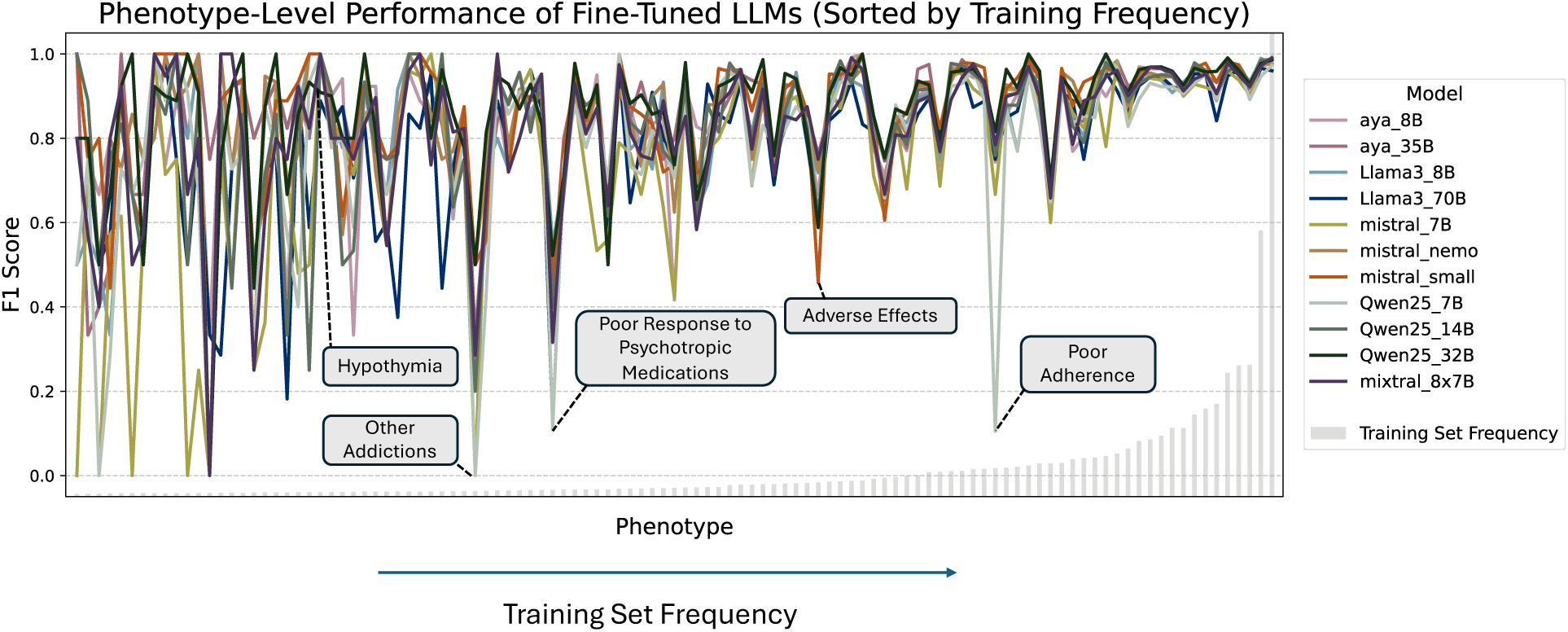
Per-phenotype performance of all fine-tuned LLMs, against frequency in the training set. Per-phenotype F1 for all LLMs fine-tuned on CSJDM data (colored lines); phenotypes are ordered by their frequency in the training data (bottom: gray bars showing the relative frequency of each phenotype in the CSJDM training set, n=1,477 documents). Highlighted phenotypes include examples of phenotypes with phenotype-specific (“other addictions”, “poor response to psychotropic medications”, “adverse effects”) and model-specific low performance (“poor adherence”), as well as high performance across-models despite low frequency in the training data (“hypothymia”).

Unlike with the base models, the tNLP method did not significantly outperform any of the fine-tuned LLMs, except for Llama3-70B (adjusted p-value < 0.0133, Supplementary Table 5). Moreover, several models - including Qwen2.5-32B, Mistral-nemo, Mistral-small, Llama3-8B, aya-8B, and aya-35B - achieved higher macro-F1 scores than the tNLP method (Supplementary Figure 3, Supplementary Table 6). Still, the tNLP method outperformed all fine-tuned LLMs in several phenotypes, including “cocaine” and “TECAR” (electro-convulsive therapy), both of which are captured by very few (2-3) distinct patterns (Supplementary Figure 5B). In contrast, all LLMs outperformed the tNLP method in the detection of several phenotypes, including “change in weight” and “altered prospection”, and were able to successfully detect concepts for which no patterns were developed due to low frequency or inter-annotator agreement (IAA) in the training data, including “psychomotor agitation” and “panic”^33^. Overall, 100 phenotypes were successfully extracted (F1>0.8) using at least one of the fine-tuned LLMs or the tNLP method.

#### Synthetic-Data Fine-Tuning

The final synthetic dataset we created to develop a publishable LLM consisted of 1,223 documents (mean number of tokens per document=83.72, SD=51.91). Application of the tNLP method to the synthetic data boosted the number of labeled instances from 3,437 to 5,427 instances (Figure 4A, Supplementary Figure 7). Given Mistral-small’s performance in both fine-tuning and few-shot settings along with its smaller size, we selected it as the base model to fine-tune on the synthetic dataset, creating Mistral-small-psych. Base Mistral-small was fine-tuned for 9 epochs, during approximately four hours on a single NVIDIA A10G GPU with 24GB of VRAM (Supplementary Note 5).

**Figure 4:**
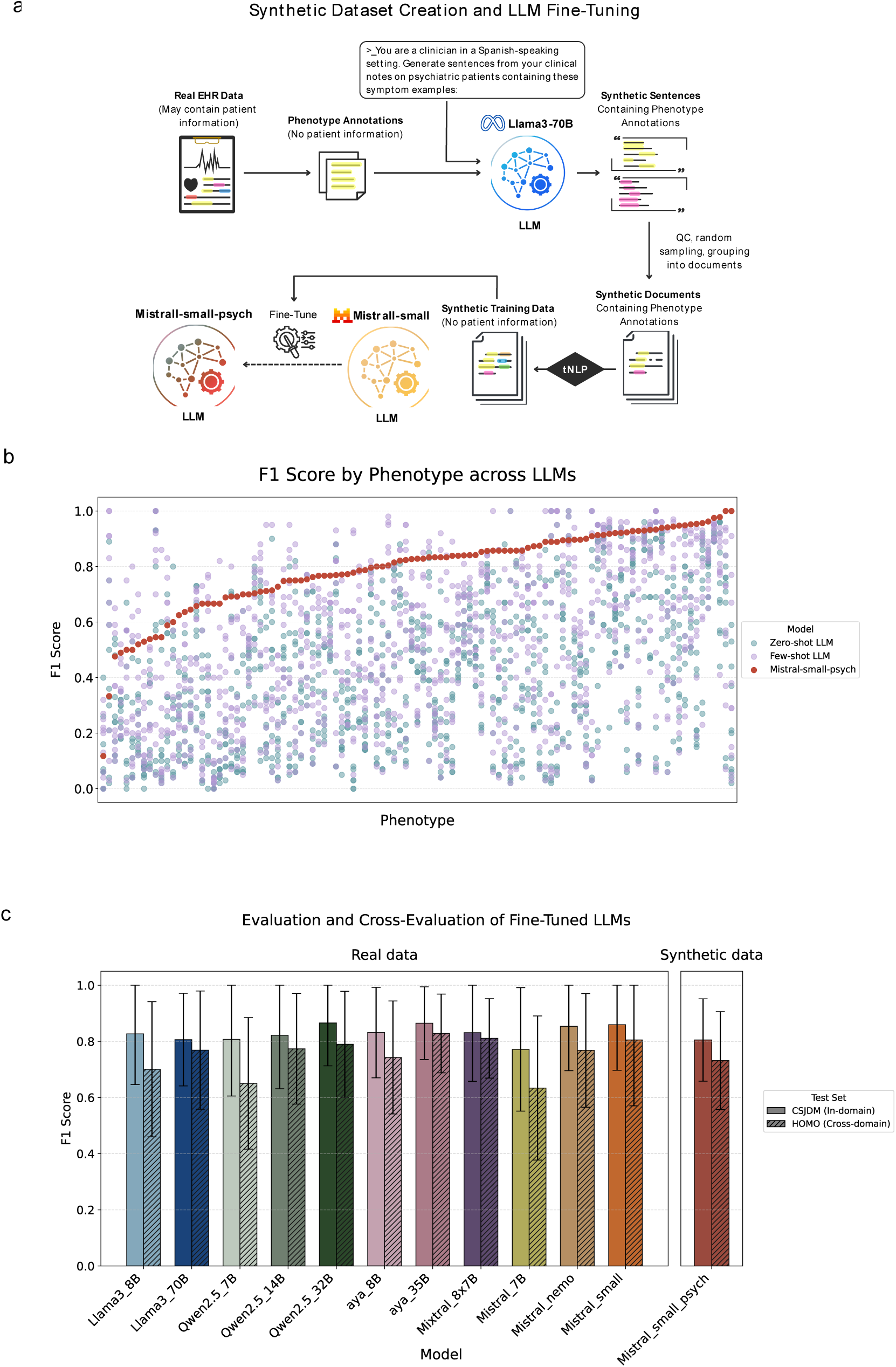
Overview of Mistral-small-psych’s creation and performance. (a) Mistral-small-psych creation: Llama3-70B was prompted to generate synthetic clinical sentences containing published phenotype annotation examples extracted from real annotated EHR data. These synthetic sentences, which contained no patient information, were sampled and assembled into documents, further labeled using the tNLP system, and used as training data to fine-tune Mistral-small, creating Mistral-small-psych. (b) Phenotype-level performance (F1) of Mistral-small-psych (red), vs. all 11 LLMs evaluated in zero-shot (blue) and few-shot (purple) settings, with phenotypes ordered by Mistral-small-psych F1 score. (c) Performance and generalizability of all fine-tuned LLMs: in-domain (CSJDM, solid bars) vs. out-of-domain (HOMO, striped bars) macro-F1 scores across all fine-tuned models. Mistral-small-psych (right, red) was fine-tuned on synthetic data. For comparison purposes, results are presented for phenotypes with three or more instances in both the CSJDM and HOMO test sets (N phenotypes=97). Error bars represent a standard deviation from the mean F1 score.

Mistral-small-psych showed good performance overall (macro-F1=0.79, mean accuracy=0.97; Table 2, Supplementary Table 4), an improvement of +0.19 from the base model’s macro-F1. Mistral-small-psych outperformed all base models in both zero- and few-shot settings (Figure 4B, Table 2), and had comparable performance to that of LLMs fine-tuned on real CSJDM data (Figure 4C). Of note, it achieved a macro-F1 score that was equal or higher than that of three models fine-tuned on real EHR data: Llama3-70B, Qwen2.5-7B, and Mistral-7B. Moreover, despite being fine-tuned exclusively on synthetic data, Mistral-small-psych achieved a mean precision of 0.82 (Table 2).

### Cross-Testing

To assess their generalizability to other settings, all fine-tuned models were also tested on the HOMO test set (n=309 documents, Figure 4C, Table 3). The fine-tuned LLMs exhibited varying degrees of out-of-domain performance, with the larger models generalizing better than the smaller ones. Fine-tuned Mixtral-8x7B showed the best generalizability, with a loss of only 0.02 (2.5%) in macro-F1, while Qwen2.5-7B had the greatest loss in performance (-0.16 points in macro-F1, or 19.75%), stemming mostly from a large drop in recall in the cross-evaluation (Supplementary Table 7).

**Table 3:** Cross-testing vs. in-domain performance results (only evaluated for concepts with support >=3 in both the CSJDM and HOMO test sets [n=97 phenotypes]).

|  | In-domain testing (CSJDM test set) |  |  |  | Out-of-domain testing (HOMO test set) |  |  |  |
| --- | --- | --- | --- | --- | --- | --- | --- | --- |
| Model | Precision | Recall | Macro-F1 | Accuracy | Precision | Recall | Macro-F1 | Accuracy |
| Llama3_70B_finetuned | 0.9 | 0.76 | 0.81 | 0.97 | 0.84 | 0.74 | 0.77 | 0.97 |
| Llama3_8B_finetuned | 0.9 | 0.79 | 0.83 | 0.98 | 0.85 | 0.64 | 0.7 | 0.97 |
| Qwen25_14B_finetuned | 0.89 | 0.8 | 0.82 | 0.98 | 0.87 | 0.74 | 0.77 | 0.97 |
| Qwen25_32B_finetuned | 0.89 | 0.86 | 0.87 | 0.98 | 0.86 | 0.76 | 0.79 | 0.98 |
| Qwen25_7B_finetuned | 0.89 | 0.76 | 0.81 | 0.97 | 0.82 | 0.58 | 0.65 | 0.96 |
| aya_35B_finetuned | 0.9 | 0.85 | 0.86 | 0.98 | 0.87 | 0.81 | 0.83 | 0.98 |
| aya_8B_finetuned | 0.88 | 0.82 | 0.83 | 0.98 | 0.81 | 0.72 | 0.74 | 0.97 |
| Mistral_7B_finetuned | 0.83 | 0.75 | 0.77 | 0.97 | 0.77 | 0.57 | 0.63 | 0.96 |
| Mistral_nemo_finetuned | 0.91 | 0.83 | 0.85 | 0.98 | 0.87 | 0.72 | 0.77 | 0.97 |
| Mistral_small_finetuned | 0.9 | 0.85 | 0.86 | 0.98 | 0.85 | 0.79 | 0.81 | 0.98 |
| Mixtral_8x7B_finetuned | 0.88 | 0.81 | 0.83 | 0.98 | 0.86 | 0.8 | 0.81 | 0.98 |
| Mistral_small_psych | 0.84 | 0.79 | 0.81 | 0.96 | 0.83 | 0.68 | 0.73 | 0.96 |

Mistral-small-psych showed moderate out-of-domain performance, surpassing the macro-F1 performance of three of the LLMs fine-tuned on real data (Llama3-8B, Qwn2.5-7B, Mistral-7B), and similar (within 0.05 range in macro-F1) to that of four LLMs fine-tuned on real data (Mistral-nemo, Qwen25-14B, Llama3-70B, aya-8B, Figure 4C).

### Synthetic Training Set Size Sensitivity Analysis

To assess the sensitivity of Mistral-small-psych’s fine-tuning process to the synthetic training data size - and the potential gains from a larger synthetic training set - we created random subsets of increasing sizes in 250 document increments. Three random subsets of each size were sampled from the original synthetic training data, and Mistral-small was fine-tuned on each training set and evaluated on the CSJDM test set.

Performance converged quickly in all aspects (mean F1, precision and recall, Supplementary Figure 8). Strikingly, a training set of only 750 documents was sufficient to reach mean recall and F1 scores of 0.77 and 0.78, nearly matching the performance achieved using the full synthetic training set. Moreover, the random samples of each size category yielded highly similar results (performance within 0.01 of the mean F1 values for each subset size), suggesting the training procedure’s robustness to varying selections of synthetic training documents.

### Error Analysis

The full phenotype-level error statistics (true positive, false positive, false negative, true negative) for zero-shot, few-shot, and fine-tuned LLMs, are shown in full in Supplementary Tables 2,3 and 4, respectively. As previously noted, most LLMs exhibited poor performance with zero-shot prompting, largely due to high false positive rates. Few-shot prompting mitigated (but did not fully solve) this issue, in most cases.

For the fine-tuned models, we observed higher performance across all fine-tuned LLMs for phenotypes with higher inter-annotator agreement (Cohen’s Kappa) and frequency in the training data (Supplementary Figure 6), in line with previous findings^33^.

The phenotypes with the greatest improvement in F1 across LLMs following fine-tuning compared with zero-shot prompting of base models were “altered emotional resonance”, “hypothymia”, “altered prospection” and “hypomanic symptoms” (all with increases of over +0.7 in average F1, Supplementary Table 4). Interestingly, most of these phenotypes had very low training set frequency (of up to 15 annotated instances in over 1,400 documents) but highly repetitive test set annotations deviating little from the phenotype name (Supplementary Table 8), emphasizing how even little training data can suffice to train the LLMs to identify these phenotypes with high sensitivity and precision.

In contrast, some broader phenotypes (“poor response to psychotropic medications”, “other addictions”, “disrobing”, “obsessions”) saw little improvement from their zero-shot performance after fine-tuning, mostly due to poor recall across models. For the most part, these phenotypes exhibited lower training and test set frequency (<20 instances in the training set) and diverse annotations in the test set, highlighting the difficulty all LLMs faced to extrapolate and capture these instances based on the limited training data. Moreover, one phenotype (“TECAR”) saw a decrease in F1 after fine-tuning compared to the base models’ zero-shot performance across all models (-0.3 F1 on average), stemming mostly from a drop in recall; this, despite all the test set annotations of “TECAR” containing the word “tecar”/“tec-ar”, in both the test set and training data (Supplementary Table 8).

While many phenotypes showed consistent gains or drops in performance across fine-tuned models, some showed low performance specific to one model (e.g. “poor adherence” for Qwen2.5-7B, or “fatigue” for Llama3-70B, Figure 3, Supplementary Table 4), indicating model-specific limitations in learning these phenotypes, rather than an issue stemming from inherent characteristics of the training or testing data.

For Mistral-small-psych, lower performance typically stemmed from low recall. We also observed a tendency to highlight spans with new phenotypes that did not appear in the training data, such as medication labels and categories (“antidepressants”), psychomotor and behavioral phenotypes (“collaboration”) and somatic phenotypes (“hypertension”, Supplementary Table 8). The number of predicted labels for these phenotypes fluctuated with training duration, peaking at 9 epochs and decreasing as training continued further (Supplementary Figure 9).

### Inference runtime, electricity consumption and cost

Two important limitations of generative AI are cost and carbon footprint. Larger models had longer runtimes and electricity consumption rates (Supplementary Table 9), with aya-35B and Llama3-70B having the longest runtime (85.62 and 81.11 seconds to phenotype 100 documents, respectively). Llama3-70B also had the highest electricity consumption (12.2 WH), nearly 7-fold that of its smaller counterpart Llama3-8B.

## Discussion

In this study, we comprehensively evaluated the ability of 11 state-of-the-art decoder-based LLMs to extract a wide range of psychiatric phenotypes from real EHR notes, assessing both “out-of-the-box” and fine-tuned performance, including external validation. Our contributions are two-fold: first, to the best of our knowledge, this is the largest study on the ability of generative AI to extract psychiatric symptoms from EHRs, both in terms of the number of models evaluated and the number of phenotypes targeted. Second, we focus on Spanish clinical text, aiming to help close the gap in NLP research and applications in non-English settings.

We further created and published an open-source generative LLM fine-tuned on synthetic documents drawing from real symptom annotations, designated to detect and extract over 100 psychiatric phenotypes from Spanish clinical text.

Our results demonstrate that zero- and few-shot model performance was generally limited due to a tendency to over-predict positives. Nevertheless, few-shot prompting yielded some improvement in both recall and precision over zero-shot performance in most cases. In addition, larger models did not necessarily yield better results overall. The limited out-of-the-box performance on this task could be attributed in part to the following two factors: first, the specificity of the task itself, as psychiatric notes contain domain-specific language and terms, as well as nuanced symptomatology; Second, the relatively small fraction of Spanish-language data in the LLMs’ pre-training corpora (for instance - for Llama3 models, less than 10% of the total pre-training data corresponded to all non-English language tokens, combined^34,44^).

Fine-tuning led to dramatic performance gains in all models, with the top-performing models (aya-35B, Qwen2.5-32B, and Mistral-small) achieving a macro-F1 score of 0.86; moreover, several fine-tuned models achieved higher macro-F1 scores compared to the tNLP method developed from the same data. In line with the literature^45,46^, these findings showcase the importance of task-specific model adaptation in enabling LLMs to perform effectively in clinical phenotyping.

As expected, fine-tuned LLMs generally performed better on phenotypes that were more common in the training data, while the detection of some phenotypes remained challenging even after fine-tuning. In most cases, these challenges appeared to be phenotype-specific (e.g. “adverse effects”), likely due to their high textual variability in the annotated documents. In other cases (e.g. Qwen2.5-7B’s performance on “poor adherence”), challenges were more pronounced for specific models, suggesting inherent limitations in certain models’ ability to learn the detection of these phenotypes. Of note, some phenotypes (e.g. “hypothymia”) had high performance across models despite low frequency in the training data, likely attributed to their simple and repetitive textual instances in the data.

Cross-evaluation of the fine-tuned LLMs demonstrated their overall generalizability, though the degree of out-of-domain performance loss varied by model and phenotype, mostly driven by changes in recall. Larger LLMs generalized better to the new setting, in line with previous findings on LLM generalization^47,48^.

We created an open-source fine-tuned generative LLM, Mistral-small-psych, for the comprehensive extraction of psychiatric phenotypes from Spanish-language clinical text and EHR documents. This privacy-preserving open-source LLM is easy to deploy and use, and can be further fine-tuned with additional annotated data for further adjustment to local documentation practices. In practice, this LLM can be applied in clinical settings for the task of symptom extraction from medical records.

Benchmarking results revealed substantial variability in runtime and cost across the evaluated LLMs, and these differences would compound when applying these models to large repositories of clinical notes. Hence, we prioritized accessibility^23^ in the creation of Mistral-small-psych by selecting Mistral-small, which yielded the smallest top-performing model upon fine-tuning on real data, as our base model. Despite being trained solely on AI-generated content, Mistral-small-psych exhibited high precision - suggesting a low incidence of hallucinations - and small performance drops in the external validation.

All fine-tuned models, including Mistral-small-psych, were fine-tuned to output not only the phenotypes detected in the input text, but also their corresponding labeled spans, precisely identifying the tokens associated with each phenotype. This feature adds a layer of explainability to the LLMs’ predictions, which is especially critical when deploying LLMs in clinical settings, where interpretability can facilitate validation and error analyses and enhance user trust^3^.

Training data size sensitivity analysis showed that fine-tuning on approximately 60% of the synthetic training data lead to nearly equal performance as fine-tuning on the full dataset, suggesting diminishing benefit from additional synthetic training data and highlighting the effectiveness of the fine-tuning process, even with relatively small (<1000 documents) fine-tuning corpora.

This study has limitations and highlights important directions for future work.

The performance and evaluation of fine-tuned LLMs inherently depend on the annotated corpus they are based on. As a result, these models are sensitive to data drifts and changes in clinical documentation practices over time. Like any machine learning system utilized in healthcare, they should be periodically re-evaluated and updated to ensure reliability.

In addition, while models were cross-evaluated to assess their transportability, this test set was also derived from a psychiatric hospital in Colombia; since Spanish is spoken across many countries with regional linguistic variations, evaluating the models’ generalizability using data from different countries would be a valuable next step.

This study presents the large-scale evaluation and development of multiple generative LLMs for comprehensive extraction of transdiagnostic psychiatric phenotypes from clinical text, as well as an open-source generative LLM specifically designed to detect these phenotypes from Spanish EHR documents. These models have broad utility across clinical and research settings, with the ultimate goal of advancing global precision psychiatry.

## Data Availability

The full annotated corpus used to evaluate and fine-tune all LLMs consists of clinical notes containing patient data, and cannot be shared publicly; however, the specific spans annotated for all concepts do not contain private health information, and were previously published in33. The synthetic dataset used for the creation of mistral-small-psych can be found at: https://github.com/clarafrydman/LLMs_for_psychiatric_phenotyping, along with the code workflow applied for inference, fine-tuning and evaluation.
Mistral-small-psych is open-source and will be made publicly available online at huggingface.co/loeslab/mistral_small_psych. The full set of patterns developed as part of the tNLP method is also publicly available at: https://github.com/clarafrydman/Spanish_Psych_Phenotyping. Further enquiries can be directed to the corresponding author.

## Compliance Statement

All procedures were performed in compliance with Colombian and United States laws and institutional guidelines and have been approved by the Institutional Review Board (IRB), Medical Institutional Review Board 3, at UCLA (IRB#20-000149, IRB#20-001537 and IRB#16-002084), the Comité de Ética del Instituto de Investigaciones Médicas at Universidad de Antioquia (UdeA), the Comité de Ética en Investigación de la E.S.E at HOMO, and the Comité de Bioética de Clínica San Juan de Dios at CSJDM. Participants provided signed informed consent to be part of studies IRB#20-000149, IRB#20-001537 and IRB#16-002084; in addition, a waiver of consent and IRB approval by Comité de Bioética de Clínica San Juan de Dios at CSJDM was granted to access all EHR data. All privacy rights of human subjects have been observed.

## Data and Code Availability

The full annotated corpus used to evaluate and fine-tune all LLMs consists of clinical notes containing patient data, and cannot be shared publicly; however, the specific spans annotated for all concepts do not contain private health information, and were previously published in^33^. The synthetic dataset used for the creation of mistral-small-psych can be found at: https://github.com/clarafrydman/LLMs_for_psychiatric_phenotyping, along with the code workflow applied for inference, fine-tuning and evaluation.

Mistral-small-psych is open-source and will be made publicly available online at huggingface.co/loeslab/mistral_small_psych. The full set of patterns developed as part of the tNLP method is also publicly available at: https://github.com/clarafrydman/Spanish_Psych_Phenotyping. Further enquiries can be directed to the corresponding author.

## Author Contributions Statement

CFG: conceptualization, methodology, analysis, writing of original draft; AA: data curation, methodology, analysis; MPV and JDLM: methodology, annotation of clinical notes; JVE: project administration; MC: methodology; AATB: conceptualization and model selection, methodology; NBF: conceptualization, methodology, supervision, writing of original draft; CLJ: methodology, data curation, supervision; LMOL: conceptualization, methodology, supervision, writing of original draft. All authors read, edited and approved the final manuscript.

## Funding Statement

This work was supported by the National Institute of Mental Health, grant numbers: R01MH123157 (to LMOL, CLJ, and NBF), R01MH137219 (to LMOL), R00MH116115 (to LMOL), and U01MH125042 (to LMOL, CLJ, and NBF), and by the UCLA Cota-Robles Fellowship (to CFG).

## Competing Interests

All authors declare no financial or non-financial competing interests.

## Notes

### Competing Interest Statement

The authors have declared no competing interest.

### Author Declarations

The Institutional Review Board (IRB) Medical Institutional Review Board 3 at UCLA (IRB#20-000149, IRB#20-001537 and IRB#16-002084) gave ethical approval for this work. The Comité de ética del Instituto de Investigaciones Médicas at Universidad de Antioquia (UdeA) gave ethical approval for this work. The Comité de ética en Investigacién de la E.S.E Hospital Mental de Antioquia gave ethical approval for this work. The Comité de Bioética de Clínica San Juan de Dios at Clínica San Juan de Dios Manizales gave ethical approval for this work. Participants provided signed informed consent to be part of studies IRB#20-000149, IRB#20-001537 and IRB#16-002084; in addition, a waiver of consent and IRB approval by Comité de Bioética de Clínica San Juan de Dios at Clínica San Juan de Dios Manizales was granted to access all EHR data. All privacy rights of human subjects have been observed. All procedures were performed in compliance with Colombian and United States laws and institutional guidelines.

