## Supplementary Figures and Notes for "Large Language Models for Psychiatric Phenotype Extraction from Electronic Health Records"

**Supplementary Material:**

**Supplementary Figures:**

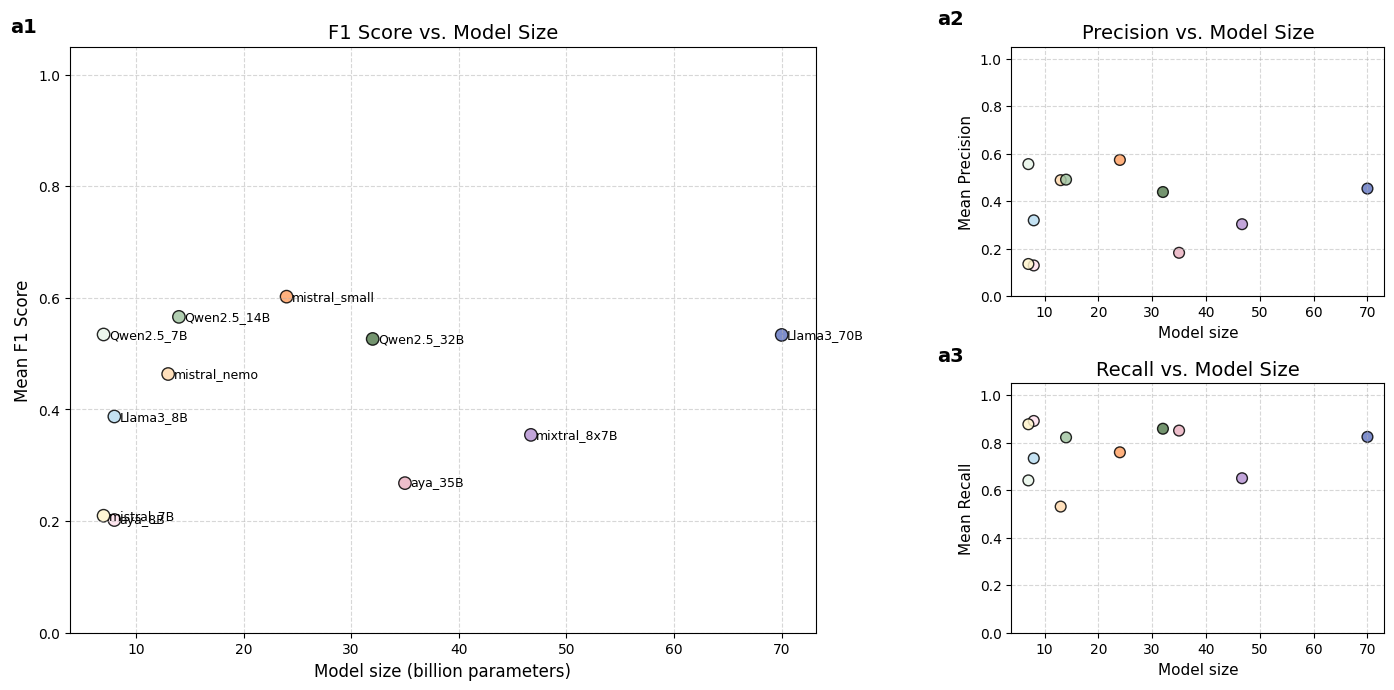

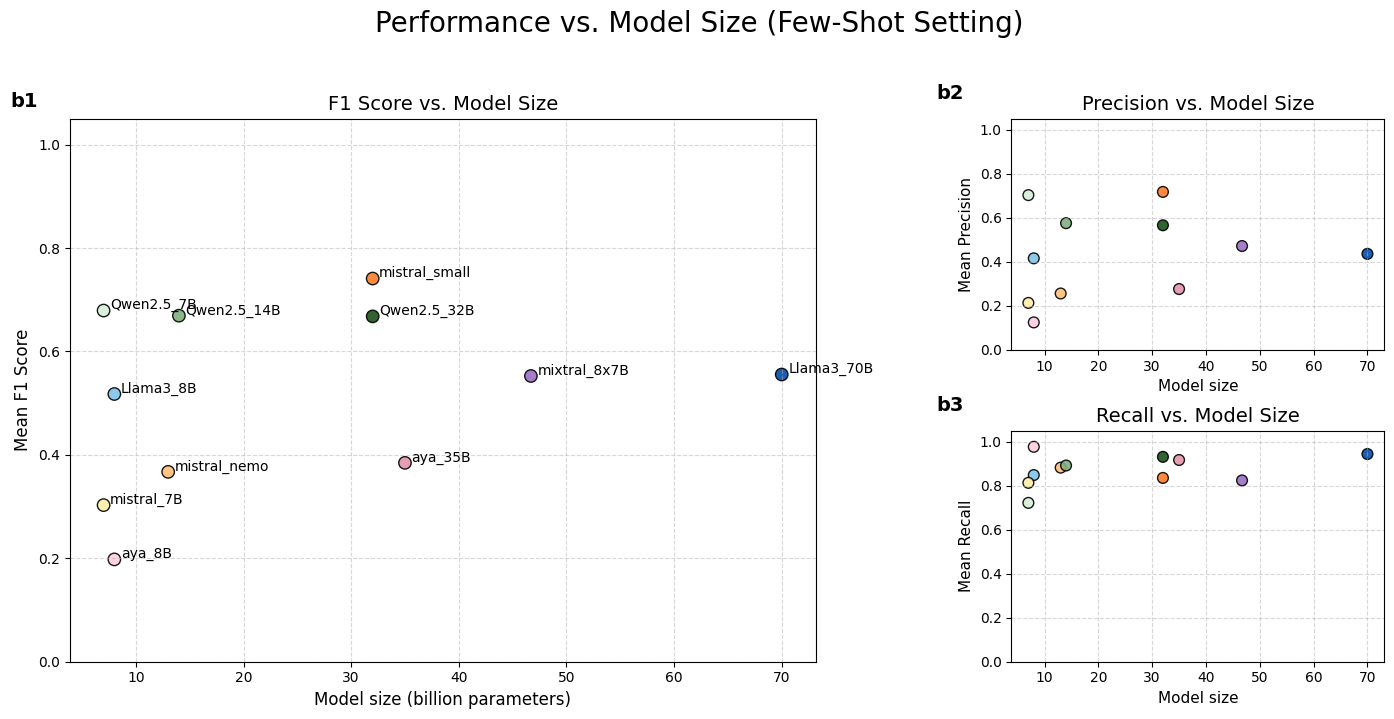

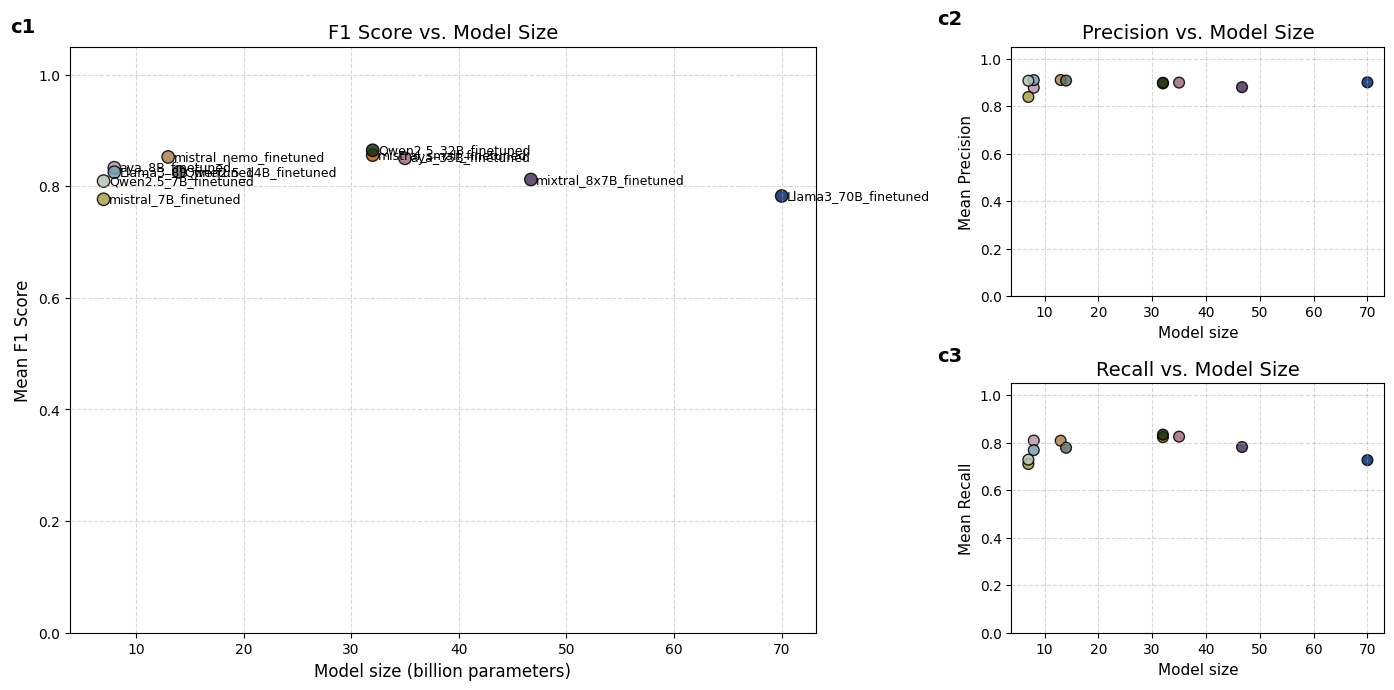

**Supplementary Fig 1:** Zero-shot (A), few-shot (B) and fine-tuned (C) performance of all LLMs as a function of model size (in billions of parameters). (1) Mean F1 score (macro-F1) across all phenotypes. (2) Mean precision. (3) Mean recall.

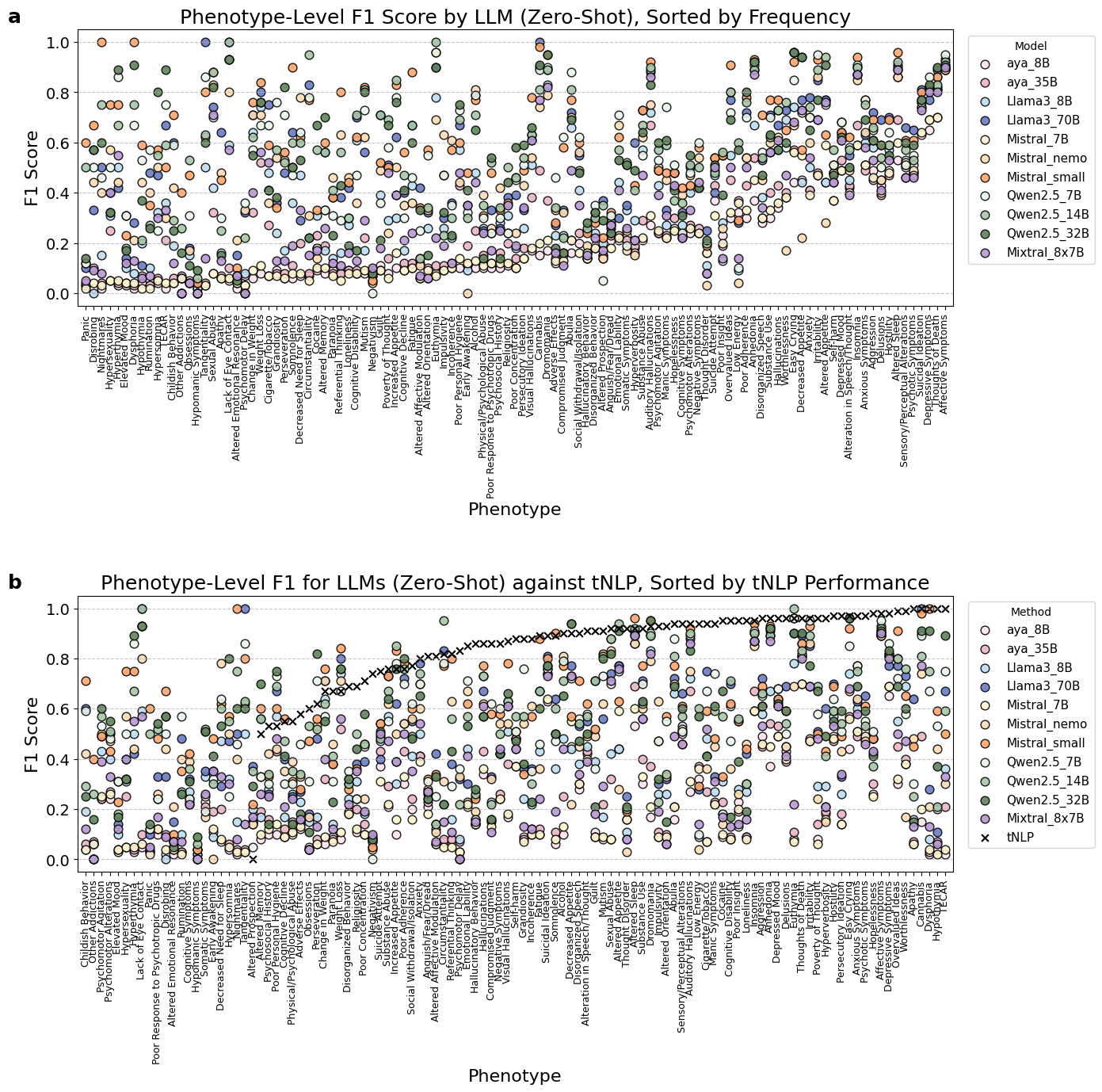

**Supplementary Figure 2**: (A) Zero-shot per-phenotype performance (F1 scores) of all LLMs, evaluated on the CSJDM test set (n=358 documents). Phenotypes are ordered by their support in the training set.

(B) Zero-shot phenotype-level F1 performance of all LLMs (colored circles) compared to the pattern-based tNLP method (black crosses) on the CSJDM test set. tNLP performance is shown only for phenotypes it was developed and evaluated on (n=88 phenotypes, out of the total 109 phenotypes shown).

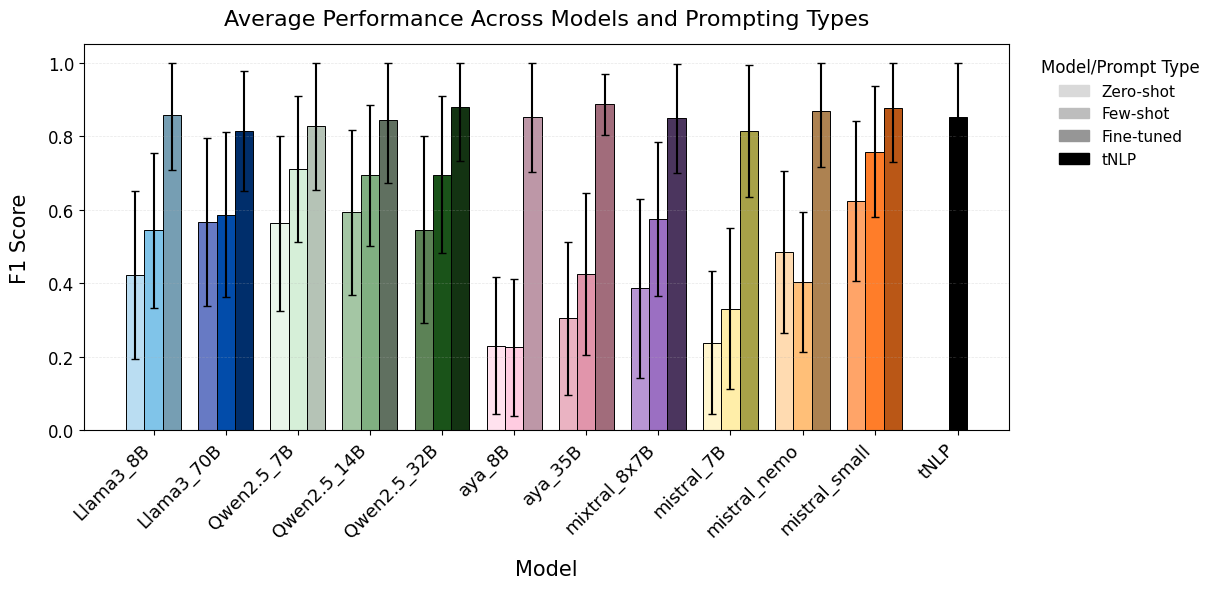

**Supplementary Figure 3:** Mean performance of zero-shot, few-shot and fine-tuned LLMs on CSJDM test set (88 phenotypes).

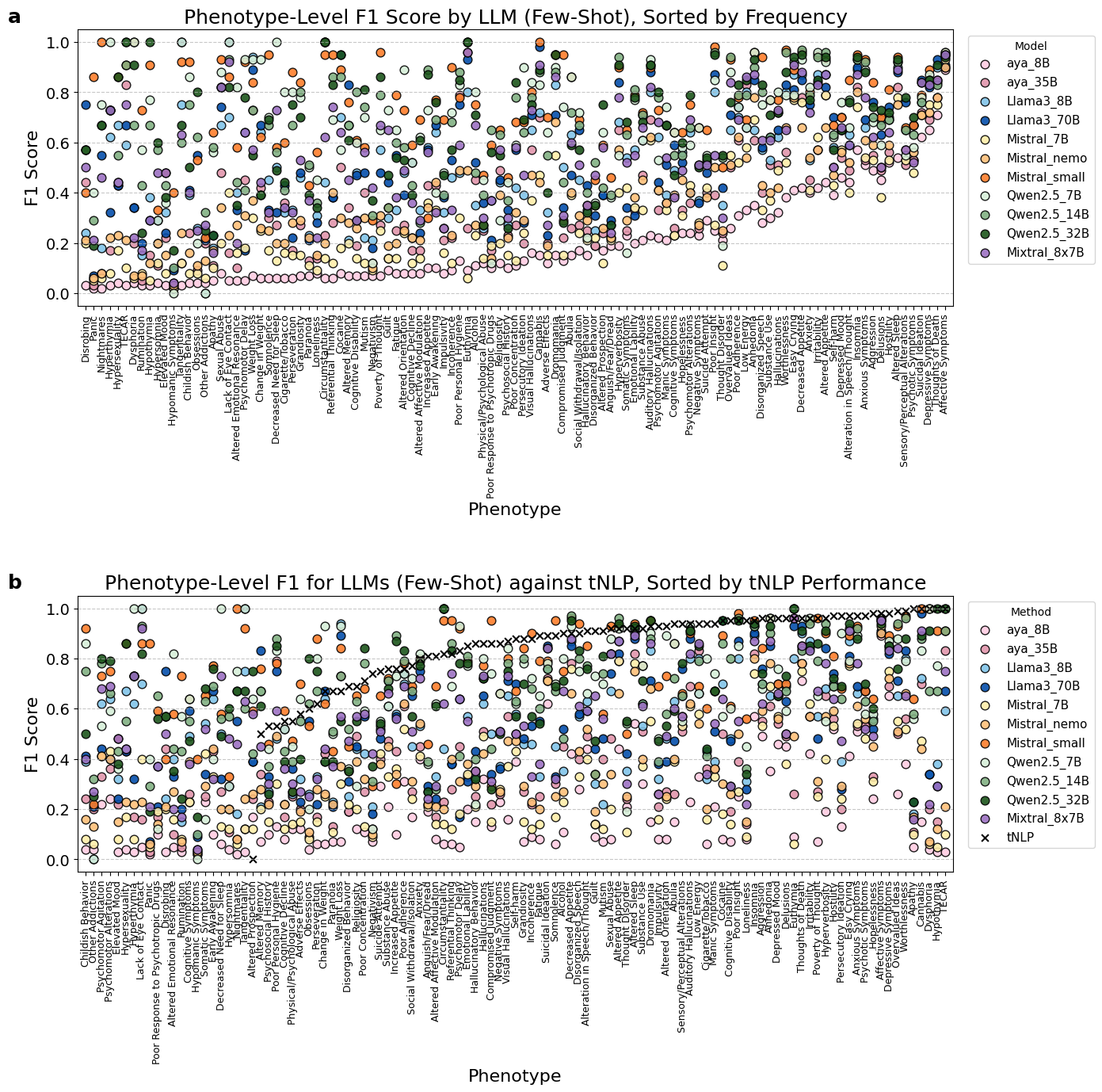

**Supplementary Figure 4**: (A) Few-shot per-phenotype performance (F1 scores) of all LLMs, evaluated on the CSJDM test set (n=358 documents). Phenotypes are ordered by their support in the training set.

(B) Few-shot phenotype-level F1 performance of all LLMs (colored circles) compared to the pattern-based tNLP method (black crosses) on the CSJDM test set. tNLP performance is shown only for phenotypes it was developed and evaluated on (n = 88 phenotypes, out of the total 109 phenotypes shown).

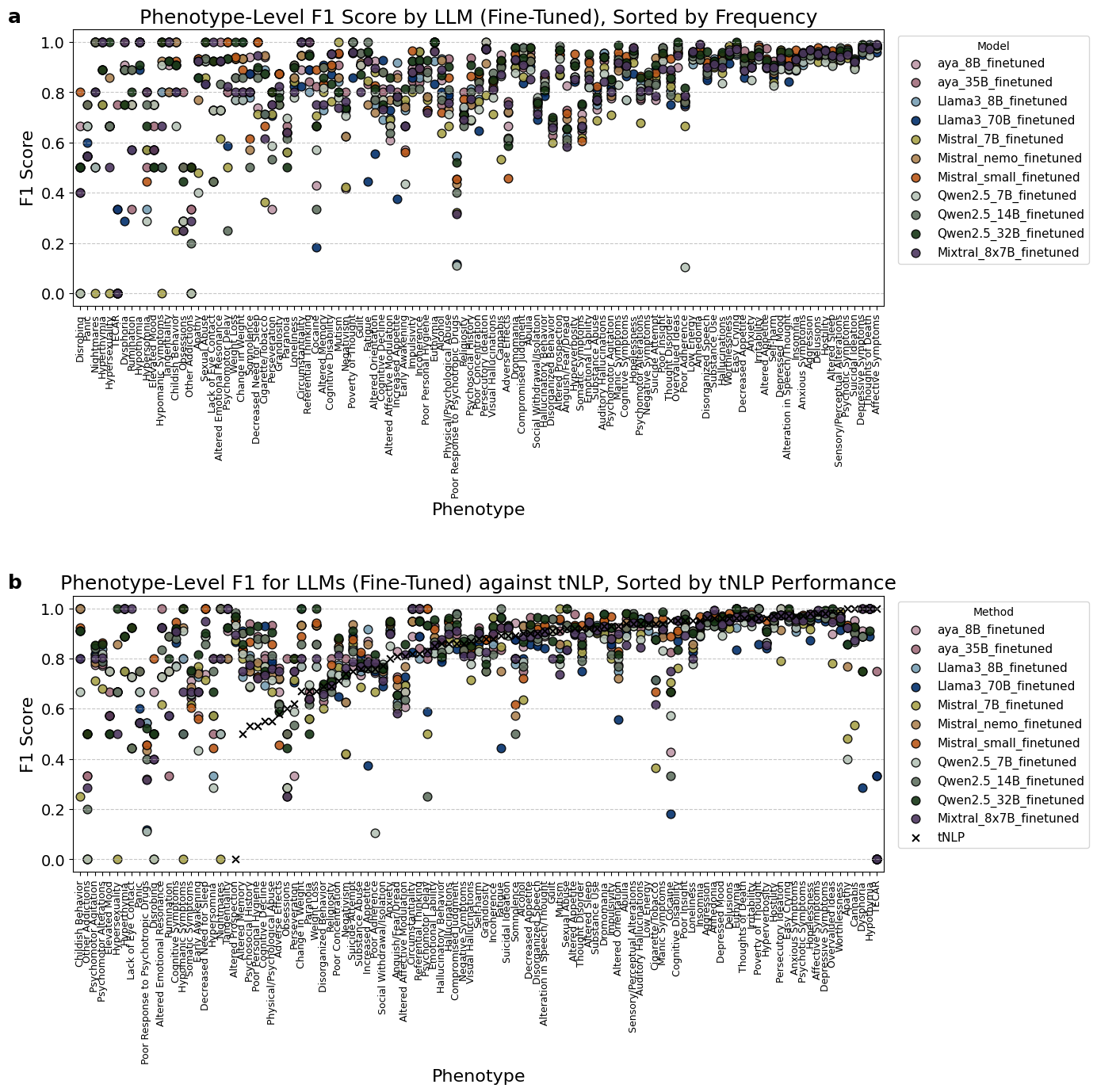

**Supplementary Figure 5**: (A) Per-phenotype performance (F1 scores) of all LLMs fine-tuned on the CSJDM training set (n=1477) and evaluated on the CSJDM test set (n=358 documents). Phenotypes are ordered by their support in the training set. (B) Phenotype-level F1 performance of all LLMs (colored circles) compared to the pattern-based tNLP method (black crosses) on the CSJDM test set. tNLP performance is shown only for phenotypes it was developed and evaluated on (n = 88 phenotypes, out of the total 109 phenotypes shown).

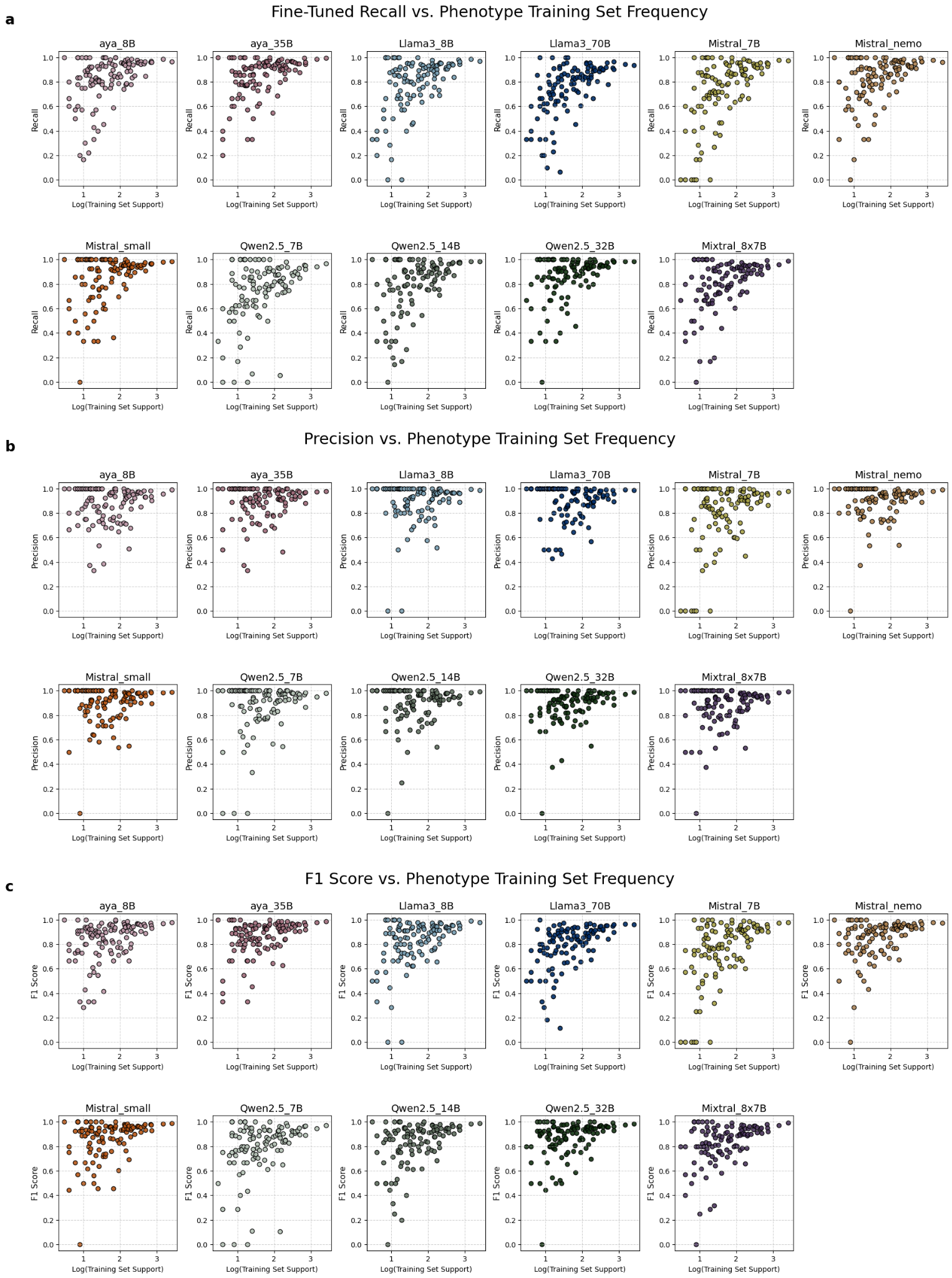

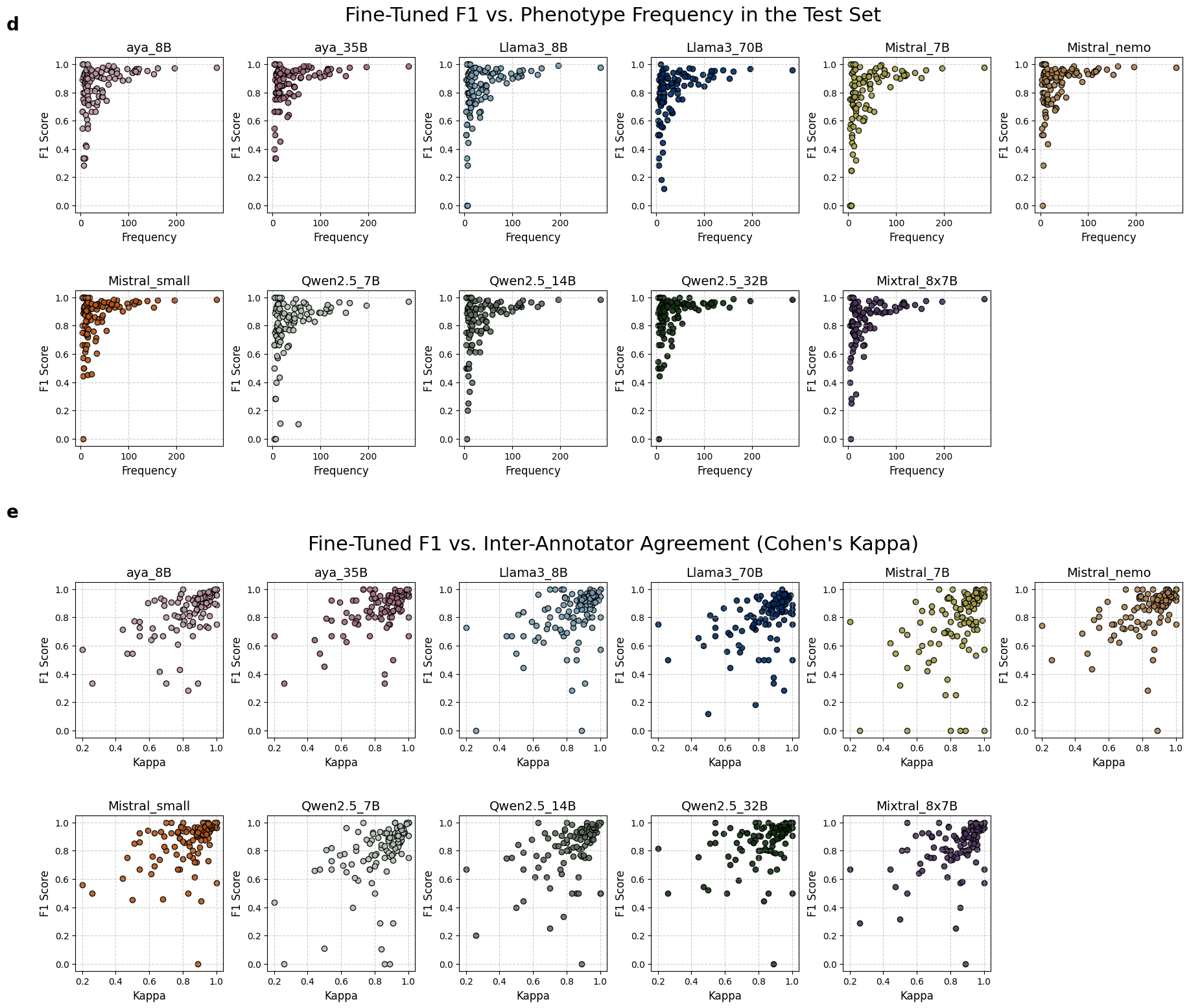

**Supplementary Fig 6:** Per-phenotype recall (A), precision (B), and F1 (C) of LLMs fine-tuned on the CSJDM training set (n = 1477) and evaluated on the CSJDM test set (n = 358), against log-transformed training set support, followed by Per-phenotype F1 against test set support (D) and phenotype inter-annotator agreement (Cohen’s Kappa, E).

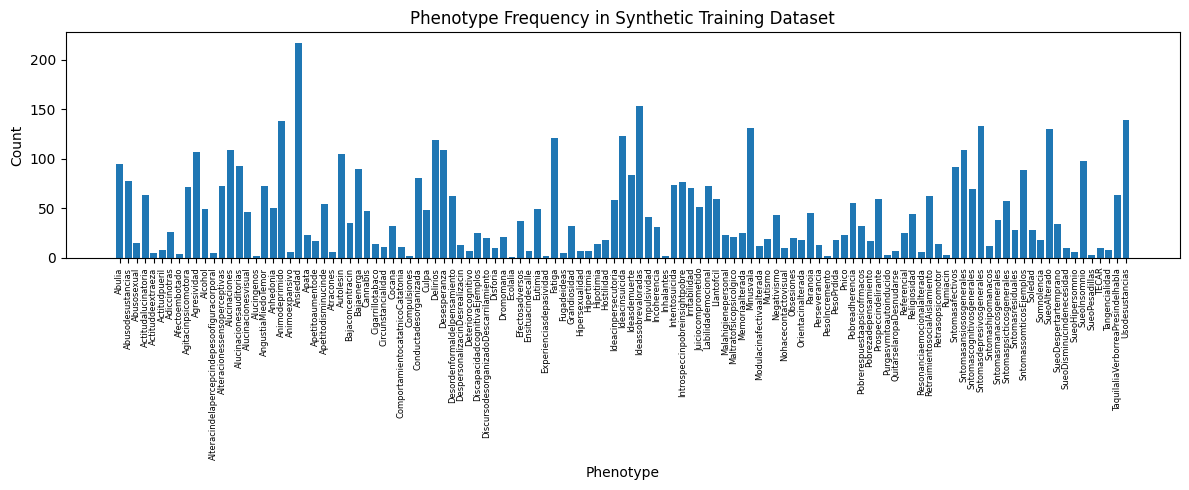

**Supplementary Figure 7**: Base phenotype frequency in the synthetic training set (n=1,223 synthetic documents).

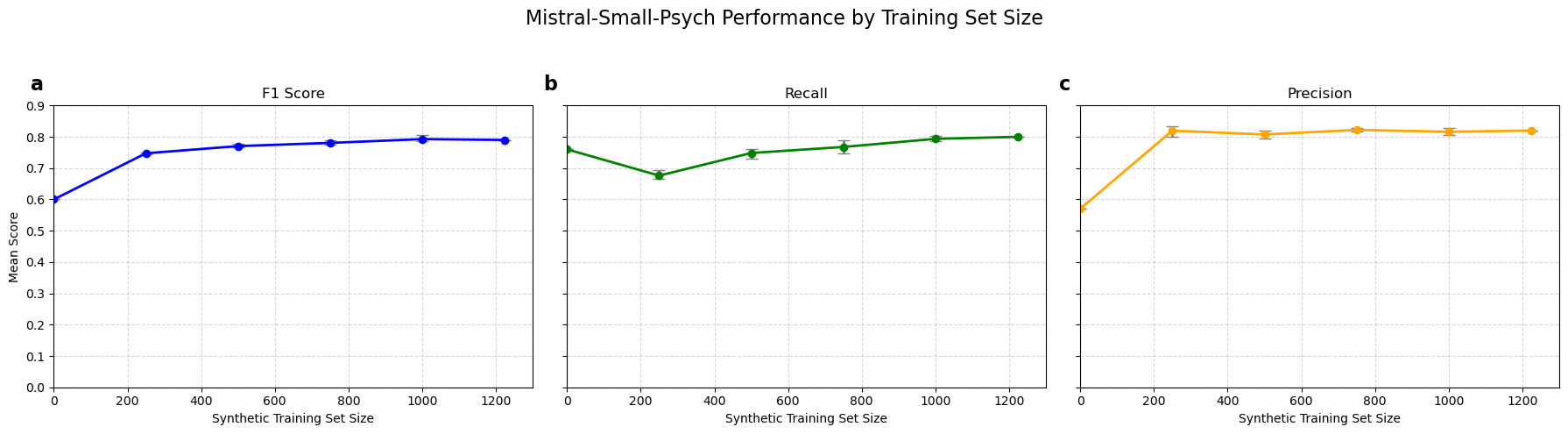

**Supplementary Figure 8:** Saturation curves showing Mistral-small-psych’s performance (y-axis) on the CSJDM test set (n=358 documents) as a function of the synthetic training set size (x-axis). Plots show mean F1 (a, blue, left), recall (b, green, center) and precision (c, yellow, right) after training on randomly sampled subsets of the synthetic documents. Subset sizes include 0 (y-axis intercept, corresponding to Mistral-small’s base zero-shot performance, defined as performance with a training set of size 0), 250, 500, 750, 1000, and the full synthetic training set (n=1,223). All points, except for sizes 0 and 1,223, represent the mean performance across three independent random samples of the same size, and the error bars represent the minimum and maximum performance values observed across the three replicates.

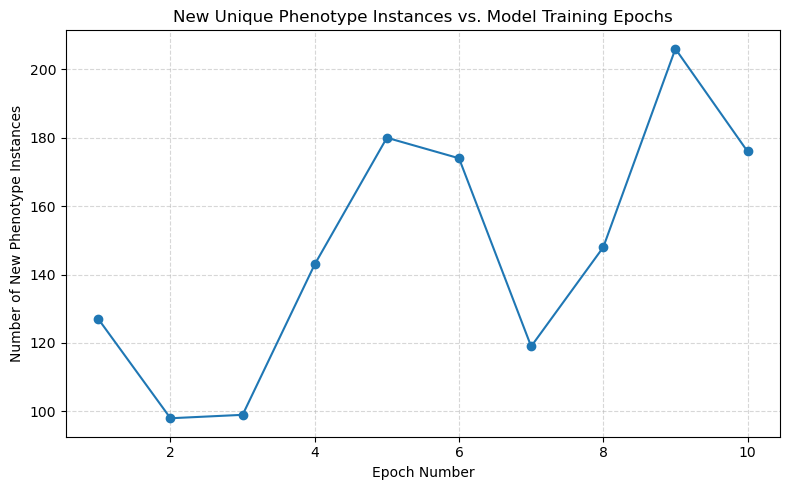

**Supplementary Figure 9:** Number of unique phenotypes labeled by Mistral-small-psych for phenotypes not in the original training data, predicted on each model training epoch.

**Supplementary Notes:**

**Supplementary Note 1: Synthetic Dataset Creation**

To create the initial pool of synthetic sentences, we utilized Llama3-70B, which was found to produce the best results among all LLMs tested in this study through an empirical comparison using different prompting strategies.

For each phenotype, we prompted Llama3-70B, providing CSJDM training set annotations (hence: “literals”), as published in^1^. In addition, to emulate the real CSJDM EHR training data, we prompted the LLM to produce sentences with no mental-health-related symptoms, but containing somatic symptoms, laboratory test results, and medical history (hence “negative sentences”). The following prompts were used to generate synthetic sentences for phenotype *P,* and negative sentences, respectively:

| Prompt | Temperature |
| --- | --- |
| “You are a Spanish-speaking clinician. Here are a few examples of mentions of the concept <P>: <annotation_example_list>. Imagine you are a Spanish-speaking clinician. Please generate at least 100 sentences from your clinical notes in Spanish about various patients with instances of the symptom <P>, each sentence containing one of these examples. You must generate at least one sentence for every example I provided. Use the examples exactly as I provided them (but without quotes). All sentences must be in Spanish, with a new line for each sentence. After each sentence, include a pipe ('\|') and then the example mention that you included in the note, exactly as I provided it.” | 0.8 |
| “Imagine you are a Spanish-speaking clinician. Please generate at least 40 [100] sentences from your clinical notes in Spanish about various patients, that do not contain any mentions of psychiatric symptoms. Sentences can contain information about the patients' age, residence, occupation, living situation, treatment plans and clinical history. Sentences should also contain information on lab test results, medications and prescriptions (name of medications and dosages), past medication history, quotes from the patient, and general somatic health conditions, but no psychiatric or mental health-related conditions." | 0.99 |

The LLM was served via Ollama, on an AWS instance with 4 A10G Tensor Core GPUs with a total of 96 GB of VRAM. Llama3-70B was prompted to generate sentences in batches of 40-100 sentences. Then, a quality control step was applied, in which numberings were removed and sentences not containing their assigned annotation literals were filtered out. Next, we randomly sampled sentences for each phenotype, prioritizing the inclusion of all distinct literals before sampling additional sentences up to reaching the frequency of the phenotype in the original CSJDM training data, if necessary. This process yielded a total of 3,440 sentences, corresponding to 119 phenotypes.

In addition, 100 sentences were randomly capitalized, imitating the style in the real CSJDM text. Then, we randomly sampled 300 negative sentences, for a total of 3,740 sentences. All sentences were shuffled and assembled into groups of 1-6 sentences, for a total of 1,223 synthetic documents.

Since the generated synthetic sentences were intended to emulate free text written about psychiatric patients, they often contained additional descriptions of psychiatric phenotypes, other than the ones explicitly prompted for. To also label these additional symptoms incidentally appearing in the synthetic dataset and create a more accurately labeled training dataset, we applied the tNLP method to the synthetic documents as a weak-labeling approach.

Finally, the dataset was converted into alpaca format, with the input being the documents and the output being the labeled spans and their corresponding phenotype labels**.**

**Supplementary Note 2: Zero-Shot Inference**

The following prompt was used to query each LLM for the detection of phenotype *P* in document *D*:

“The following is a document in Spanish for a controlled and safe analysis task. Respond '1' if this text contains an explicit mention or instance of the concept: <P>, and '0' otherwise. Respond '1' even if the symptom appears in a negated, hypothetical, or historical context, or described a different person. Do not assume the presence of the symptom if it does not explicitly appear in the note in a mention or example. Reply just '1' or '0', nothing more. The note is: <D>”

No additional information was provided to the models, and no context was carried across queries. Responses given in the wrong format (most often general descriptions and reiterations of the document or statement that the document does not contain an explicit mention of the document, e.g.: “The note does not contain an explicit mention or instance of the concept ‘Abulia’”) were classified as negative responses. For higher reproducibility and output stability, all evaluations were done under temperature=0.

**Supplementary Note 3: Few-Shot Inference**

For few-shot prompting, up to five distinct annotations of each phenotype were provided to the LLMs as examples in the prompt, along with the document. All examples were selected from the CSJDM training set. Selecting the most common annotated spans in the training set often results in highly similar spans (for example, for the phenotype “psychotic symptoms”, the most common annotations were “SÍNTOMAS PSICÓTICOS”, “sintomas psicóticos”, and “SINTOMAS PSICOTICOS” – all highly similar spans, introducing little variation and no alternative phrasing). Therefore, to balance representativeness and diversity, phenotype annotation examples were selected using the following strategy: first, we ranked all annotated instances in the training set by frequency, and then randomly selected five examples from the top ten most common different annotations. If no more than ten or five different annotations were annotated for the phenotype, we selected all instances to randomly sample from, or all different annotated instances to construct the few-shot prompt, respectively. This ensured that we would include common annotations as few-shot examples, providing power to the prompt, while also introducing some diversity. For each phenotype, the same examples were provided when prompting for each test set document.

The following prompt was used to query each LLM for the detection of phenotype *P* in document *D,* given annotated example list *E*:

"I will provide a document in Spanish for a controlled and safe analysis task.

Your task is to respond '1' if this note contains an explicit mention or instance of the concept: <P>, and '0' otherwise. Respond '1' even if the symptom appears in a negated, hypothetical, or historical context, or is described for a different person. Do not assume the presence of the symptom if it does not explicitly appear in the note in a mention or example. Reply just '1' or '0', nothing more. Here are a few examples of mentions of the concept <P>: <E>. And here is the document: <D>"

As in zero-shot prompting, no context was carried across queries, and responses not addressing the task (reiterations of the document’s content or explanations of why the document did not contain a given phenotype) were classified as negative responses.

**Supplementary Note 4: Fine-Tuning on Real CSJDM Data**

Models were fine-tuned using a training set consisting of 1,477 documents and a validation set of 165 documents, used to fine-tune hyperparameters. Fine-tuning was carried out on g5.2xlarge (all Mistral and Qwen2.5 models, Llama3-8B, and aya-8B) or g5.12xlarge (Llama3-70B, aya-35B and Mixtral-8x7B) AWS instances, with 1/4 A10 NVIDIA Tensor Core GPUs and 24/96 GB of VRAM, respectively. Base models were downloaded in 4-bit quantization and fine-tuned using LoRA (with r=8, alpha=16 and dropout=0.05), targeting all model linear layers. Brain Floating-Point 16-bit (Bf16) precision was used. When utilizing multi-GPU instances, models were sharded across GPUs using Fully Sharded Data Parallel (FSDP). Unsloth was utilized to reduce memory consumption on single-GPU settings for all Mistral and Qwen2.5 models, as well as Llama3-8B. Axolotl was used for the fine-tuning of all other LLMs.

Training and validation data was converted to alpaca format, as follows:

| **Alpaca prompt** | Below is an instruction that describes a task, paired with an input that provides further context. Write a response that appropriately completes the request. |
| --- | --- |
| **Instruction** | ### Instruction:  I will provide a clinical note in Spanish. Identify a list of all symptoms from the input text, even if the symptom appears in a negated, hypothetical, or historical context, or described a different person. Format your answer as a list of the phenotypes separated by new line characters and do not generate random answers. Only output the list. |
| **Input** | ### Input:  <DOCUMENT_BODY> |
| **Output** | ### Response:  <PHENOTYPE_1_TEXT>\| <PHENOTYPE_1_LABEL> <PHENOTYPE_2_TEXT>\| <PHENOTYPE_2_LABEL> <PHENOTYPE_3_TEXT>\| <PHENOTYPE_3_LABEL>  …  END<\|im_end\|>' |

Models were trained with mostly default parameters, for 1-4 epochs. For all models, we tested the following learning rates: 1e-3, 3e-4, 1e-4, 1e-5, as well as simulating a larger batch size using gradient accumulation steps=4. Following are the final training details for all models:

| **Model** | **Learning rate** | **Num epochs** | **Gradient accumulation steps** | **Training Time (Minutes)** | **Instance VRAM (GB)** |
| --- | --- | --- | --- | --- | --- |
| Llama3-8B | 3e-4 | 2 | 4 | 39 | 24 |
| Mistral-7B | 1e-5 | 3 | 1 | 81 | 24 |
| Mistral-nemo | 1e-4 | 2 | 1 | 52 | 24 |
| Mistral-small | 1e-4 | 2 | 4 | 98 | 24 |
| Qwen2.5-7B | 3e-4 | 2 | 4 | 36 | 24 |
| Qwen2.5-14B | 3e-4 | 2 | 4 | 92 | 24 |
| Qwen2.5-32B | 3e-4 | 2 | 4 | 116 | 24 |
| Aya-8B | 1e-3 | 2 | 4 | 46 | 24 |
| Aya-35B | 3e-4 | 4 | 4 | 178 | 96 |
| Llama3-70B | 3e-4 | 2 | 4 | 581 | 96 |
| Mixtral-8x7B | 3e-4 | 3 | 4 | 196 | 96 |

**Supplementary Note 5: Fine-Tuning on Synthetic Data**

To create Mistral-small-psych, Mistral-small was fine-tuned for 9 [tested 1-14] epochs on the synthetic dataset documents (n=1,223), on an AWS g5.2xlarge instance on a A10 NVIDIA Tensor Core GPU with 24 GB of VRAM. Documents were converted to the alpaca format for training (see **Supplementary Note 4** for template). The base model was fine-tuned using a learning rate of 3e-4 and a larger batch size was simulated using a gradient accumulation step=16. As with the fine-tuning on real data, the base LLM was downloaded in 4-bit quantization and fine-tuned using LoRA (r=8, alpha=16 and dropout=0.05) targeting all model linear layers and using Bf16. Unsloth was utilized to reduce memory consumption and training time. Total training time (for 9 epochs) was 4 hours.

**Supplementary Note 6: Evaluation Metrics**

Performance at all levels (for both inference and fine-tuned models) was evaluated on the CSJDM test set against the clinicians’ annotations, using precision, recall, macro-F1, and accuracy, defined as:

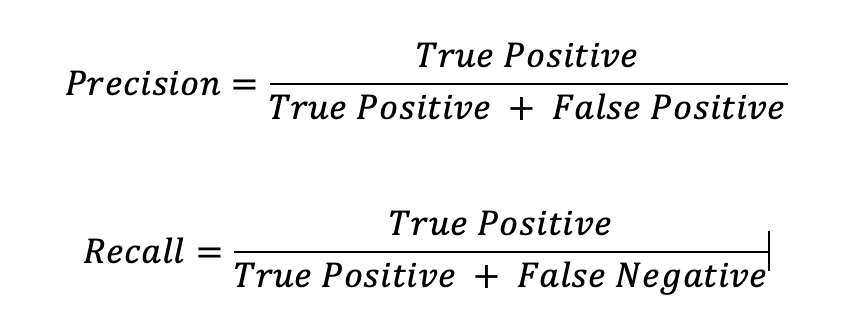

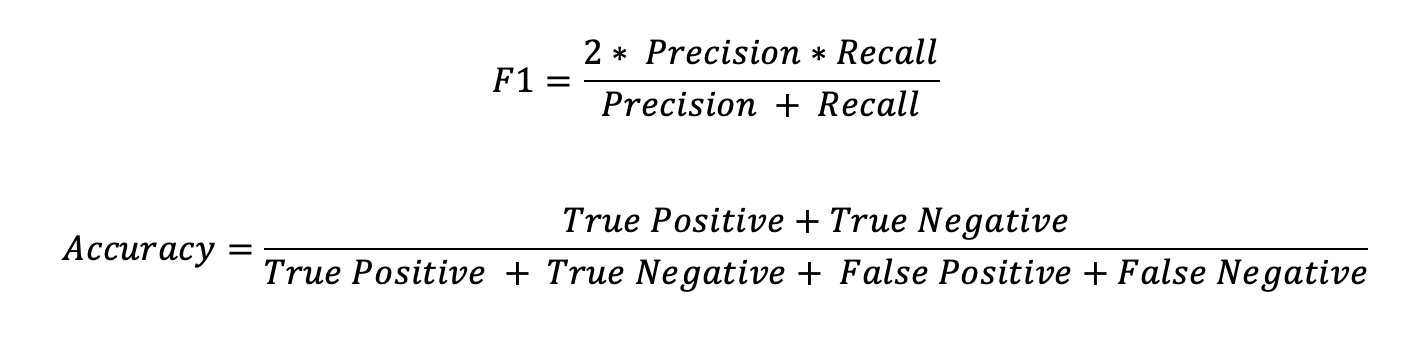
